# Remission of Type 2 Diabetes in Linked Health Records: Uncommon, Short-Lived, and More Often Ended by Treatment Restart Than by Rising HbA1c

**DOI:** 10.64898/2026.10.14.26365480

**Authors:** Felipe Lobelo, Jared Durieux, Daniel Kim, Christos Colovos

**Affiliations:** Emory Global Diabetes Research Center, Rollins School of Public Health, Emory University, Atlanta, GA; Evidence Architecture, Stow, OH; Division of Cardiology, Department of Medicine, University of Washington, Seattle, WA; Meta Platforms, Inc, Menlo Park, CA

**Author notes:** **Corresponding author**: Felipe Lobelo, MD, PhD, Emory Global Diabetes Research Center, Rollins School of Public Health, Emory University, 1518 Clifton Road NE, Atlanta, GA 30322.

## Abstract

**Objective:** To estimate how often remission of type 2 diabetes is recorded in routine care, how long it lasts, and what ends it, separating a return of hyperglycemia from a restart of treatment.

**Research design and methods:** We studied 136,781 adults aged 18 years or older with laboratory-confirmed type 2 diabetes in the Protégé linked electronic health record (EHR) and claims database. The 96,817 adults with glucose-lowering treatment recorded at or before cohort entry formed the primary (previously treated) cohort; 39,964 adults with no treatment recorded at entry were analyzed in parallel. Remission required two HbA1c values <6.5%, 90–365 days apart, the first at least 3 months after all glucose-lowering drugs were discontinued, with no therapy recorded between them. After remission we followed glycemic relapse (HbA1c ≥6.5%) and treatment restart (a new outpatient prescription or dispensed fill) separately, with death as a competing event (Aalen–Johansen).

**Results:** Remission was documented in 4.33% of previously treated adults cohort and 15.57% in the cohort of untreated adults at entry. Five-year cumulative incidence (Aalen–Johansen) was 3.82% (95% CI 3.69–3.93) and 13.27% (12.90–13.59) in each cohort, respectively. Remission was less common with greater baseline treatment burden: exploratory adjusted 5-year incidence was 4.65% in adults with 1 glucose-lowering class with a treatment episode recorded at or before entry, 3.62% with two, 2.69% with three, and 2.02% with four or more such drugs.. The Median Kaplan–Meier duration of first remission was 226 days (IQR 93–855) in previously treated adults and 371 days (166–1,027) in adults untreated at entry; 37.9% (treated cohort) and 23.1% (untreated cohort) of episodes had ended by 152 days. Among remission episodes that had ended, median duration was 159 days (71–362) and 265 days (129–565) for the treated and untreated at entry cohort, respectively. Within 5 years of remission in previously treated adults, the first event ending remission was glycemic relapse in 31.94%, treatment restart in 47.83%, and death in 4.65%.Treatment restart was about 1.5 times as common as glycemic relapse as the first event ending remission. A second remission followed within 5 years in 19.53% of previously treated and 24.10% of untreated remitters.

**Conclusions:** Recorded remission was uncommon and short-lived, and it ended more often with a treatment restart than with a rise in HbA1c. Studies of remission in real-world data should report relapse and restart separately and state how they confirm remission and establish that treatment stopped.

**Article highlights:** **Why did we undertake this study?** Most real-world studies count any end of remission as relapse, mixing rising HbA1c with treatment restart.

**What is the specific question we wanted to answer?** How often is remission recorded in routine care, how long does it last, and what ends it?

**What did we find?** About 1 in 23 previously treated adults had remission (4.33% ever recorded; 3.82% by 5 years); Remission episodes had a median duration of about 7.5 months (37.9% had ended by 5 months) and ended more often with a treatment restart than with rising HbA1c (47.83% vs 31.94% as first event by 5 years). Remission was less common with greater baseline treatment burden after exploratory adjustment (4.65% with 1 glucose-lowering class with a treatment episode recorded at or before entry, 3.62% with two, 2.69% with three, and 2.02% with four or more such drugs).

**What are the implications of our findings?** Remission studies and quality measures should report glycemic relapse and treatment restart separately.

## Introduction

Remission of type 2 diabetes now has a working definition: an HbA1c below 6.5% measured at least 3 months after glucose-lowering drugs are stopped (1). Trials of intensive weight management and of metabolic surgery showed that remission can be achieved and, with support, maintained for several years (2–8); the National Health Service in England has offered remission programs at scale (9); and remission has been linked to lower rates of kidney and cardiovascular disease (10). Much less is known about remission as it appears in routine care: how often it is recorded, how long it lasts, and what ends it.

Real-world estimates differ more than tenfold, and the differences appear to reflect how remission is defined more than the population studied. Kaiser Permanente reported a 7-year incidence of 0.14% to 1.47% with 12-month medication-free definitions (11), and a U.S. claims and laboratory study found 3.2% based on a single HbA1c (12). A study in Scotland reported a point prevalence of 4.8% (13), one in Hong Kong found 6.1% among newly diagnosed adults (14), and one in England estimated 9.7 per 1,000 person-years with HbA1c values at least 182 days apart (15). After GLP-1 receptor agonist initiation, estimates ranged from 5.8% to 18.3% depending on whether ongoing treatment was allowed (16). Recent reviews have called for clearer and more patient-centered definitions (17,18).

Two methodological gaps stand out in this literature. First, most studies record a single “relapse” that combines two different events: HbA1c rising back to the diabetes range, and a clinician restarting glucose-lowering treatment. For example, a report from six U.S. health systems found 2.9% remission over 3 years, defined by two consecutive HbA1c values below 6.5% at least 90 days apart with no medication supply recorded in between, and counted either an HbA1c of 6.5% or higher or a new medication fill as relapse (19). These events reflect different processes. A rise in HbA1c is a change in glycemic control. A treatment restart is a clinical decision, which may be prompted by a measured glucose value in the diabetes range while HbA1c is still below 6.5%, by the need for kidney or cardiovascular protection (20), or by a hospital stay that interrupted treatment. Combining the two makes remission look less durable glycemically than it may be and obscures how often, and why, each event ends remission. Second, with GLP-1 receptor agonists and SGLT2 inhibitors in wide use, an HbA1c below 6.5% while on treatment is common and clinically welcome but is not remission (1,21,22). Distinguishing these states requires tracking glycemia and treatment separately.

We used a linked dataset of 4.95 million patients to follow 136,781 adults with laboratory-confirmed type 2 diabetes (96,817 previously treated; 39,964 untreated at entry), We studied the following: (1) how often remission was recorded and how long it lasted, (2) whether remission ended through a rise in HbA1c or a treatment restart, and (3) the extent to which estimates on the frequency of remission, glycemic relapses and treatment restarts are driven by the various operational definitions that have been previously proposed

## Research design and methods

### Data source

This retrospective study used Protégé (June 2026 release), a de-identified, longitudinal database that combines electronic health records (EHR) with open medical and pharmacy claims linked to a national aggregator (pharmacy and medical claims) and to a death file compiled from Social Security verification, obituary, memorial, and claims sources. Protégé draws on routine-care EHR data from 21 anonymous data-owner values in the delivered data (the vendor attributes 33 contributing systems from a 39-system upstream network). Subsets of patients are also linked to radiology data and to dated records of death. The data release we had access to contains 4,950,000 patients residing in all 50 US states and the District of Columbia. The database was built from aggregating patients with at least five clinical notes and records linkable to open claims. Patients linked to radiology data were included preferentially.

The remaining sample was enriched for diagnostic diversity by preferentially oversampling low-prevalence clinical phenotypes within the ICD-10 hierarchy. The database is therefore not a probability sample of the US population or of the contributing health systems’ patients.

Structured EHR data include demographics; encounters in ambulatory, telehealth, emergency, inpatient, and observation settings; diagnoses; procedures (CPT, HCPCS, ICD-10-PCS, and local codes); laboratory results; vital signs; medication orders and fills; and surgical cases.

Clinical notes are de-identified, with names of people and places replaced by synthetic surrogates. Claims data include medical claim lines and headers with diagnosis and procedure codes and payer type, and pharmacy claims with National Drug Codes, days supply, and dispensing status; 99.4% of patients have linked medical or pharmacy claims. The data capture care documented by contributing health systems and services billed in open claims. Insurance enrollment periods are not available, so periods without recorded care cannot be distinguished from care received elsewhere. Cost information is limited to billed charges, mortality is all-cause, and facility identifiers are not included. Data were de-identified before delivery. Birth dates are reduced to the year, ages of 89 years and older are top-coded, residence is reported as state and 3-digit ZIP code, and names of people and places in clinical text are replaced with synthetic surrogates. Dates are shifted by a constant offset for each patient, so intervals within a patient are exact but calendar dates are approximate. For this reason, we do not report calendar dates of cohort entry or follow-up; HbA1c follow-up ended on 30 April 2025 on the shifted timeline, and calendar periods are used only as approximate descriptive groupings. Prescriptions recorded in the EHR were available throughout follow-up, whereas pharmacy claims covered 2017 through 2022. Follow-up depended on clinical contact rather than scheduled testing (23), and EHR data required explicit rules for completeness and plausibility before analysis (24,25). Reporting follows the RECORD statement (26).Before delivery data underwent deidentification by Expert Determination. An external review by Bluebonnet Ethical Review determined that this activity is appropriately classified as Not Human Subjects Research (NHSR) and that IRB review is not required. This study was supported through using an internal Meta scaffold around Muse Spark 1.3 and Wonder which assisted the authors in all the stages of the study. The model was used to draft the analyses, study cohorts, statistical plan, protocols, and during manuscript preparation. An initial protocol and statistical analysis plan were recorded, and amendments to the medication definition and the death-record checks were dated. Analyses added after the initial lock, including race × treatment, weight audits, and GLP-1 period analyses, are labelled exploratory.

### Development and confirmation samples

Remission was defined according to the ADA consensus statement (1). Applying this definition to routine records required operational rules that the consensus does not specify, including which records show that a drug was stopped or restarted, how gaps between prescriptions are bridged, how duplicate laboratory results are handled, and which death records are credible. To keep these rules from being tuned to the results, we developed and tested them in a random one-sixteenth of patients and then fixed them; this development sample was excluded from all estimates.

### Study population

Of 685,303 patients with a type 2 diabetes diagnosis code, 180,894 had a qualifying pair of HbA1c values operationalising the ADA HbA1c threshold (27): pairs were anchored on the first high HbA1c of a run (the first value of 6.5% or higher after a lower value or no prior value), with a second qualifying value 90–365 days later and no lower value in between (Fig. 1). Code lists and algorithms are in Supplementary Tables 1 and 1A. Cohort entry was the date of the second HbA1c value. We excluded patients with type 1, secondary, or monogenic diabetes; gestational diabetes or pregnancy; dialysis or kidney failure; active cancer; post-transplant status; or a first HbA1c recorded during steroid treatment, leaving a cohort of 136,781 adults (Fig. 1; Table 1).

**Figure 1.**
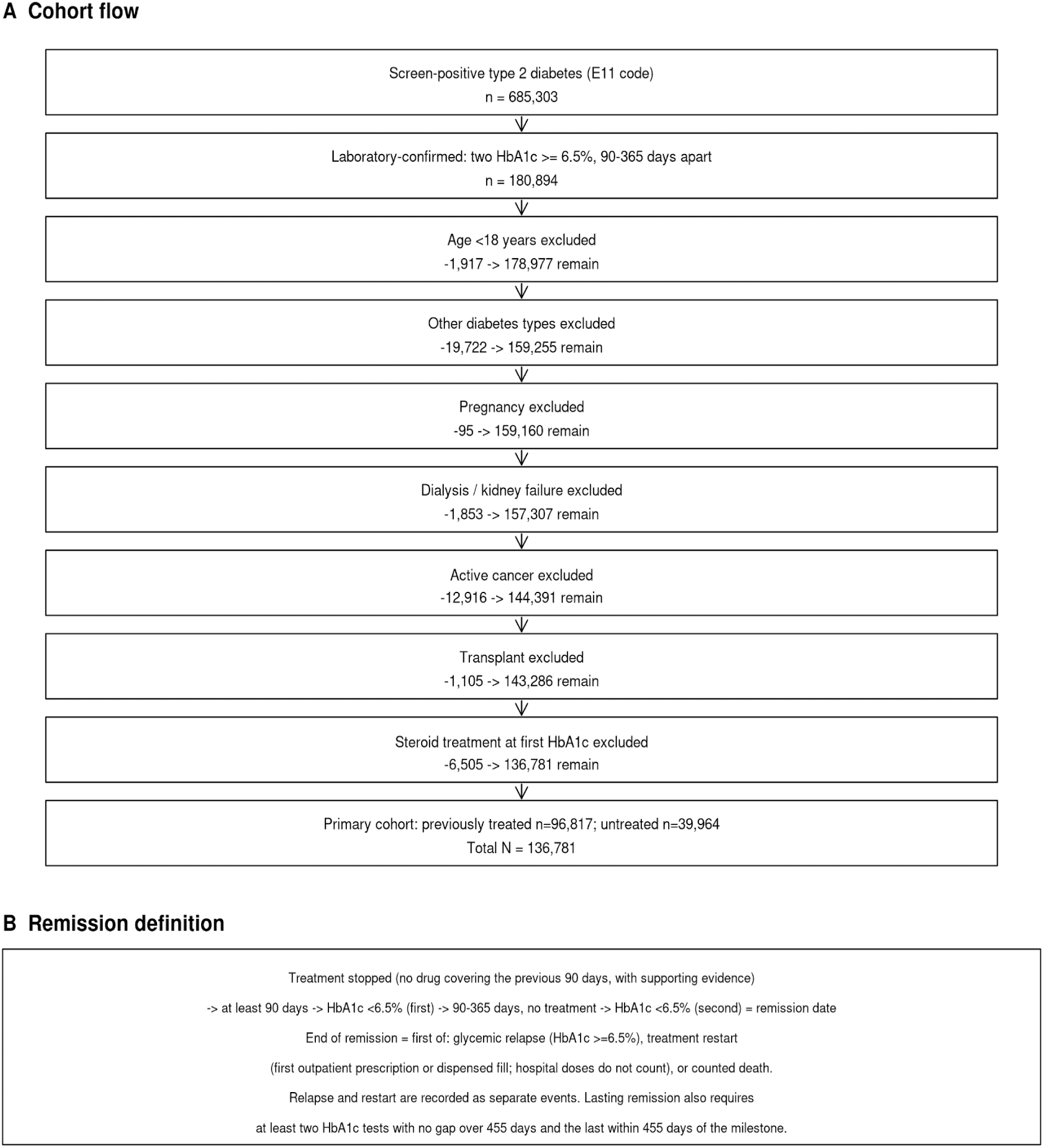
Study cohort and definitions of remission, glycemic relapse, and treatment restart in a linked EHR and claims cohort (**A:** Selection of adults with laboratory-confirmed type 2 diabetes (qualifying pair operationalising the ADA HbA1c threshold, anchored on the first high HbA1c of a run — the first value ≥6.5% after a lower value or no prior value — with a second qualifying value 90–365 days later and no lower value in between) and the sequential exclusions **B:** Remission required two HbA1c values <6.5% recorded 90–365 days apart, the first at least 90 days after all glucose-lowering drugs were stopped, with no glucose-lowering drug recorded between them; the remission date was the date of the second value. Remission ended at the first of glycemic relapse (first HbA1c ≥6.5%), treatment restart (first outpatient prescription or dispensed fill of any glucose-lowering drug; hospital doses do not count), or counted death, and continued if none of these occurred. Relapse and restart were recorded as separate events.

**Table 1.**
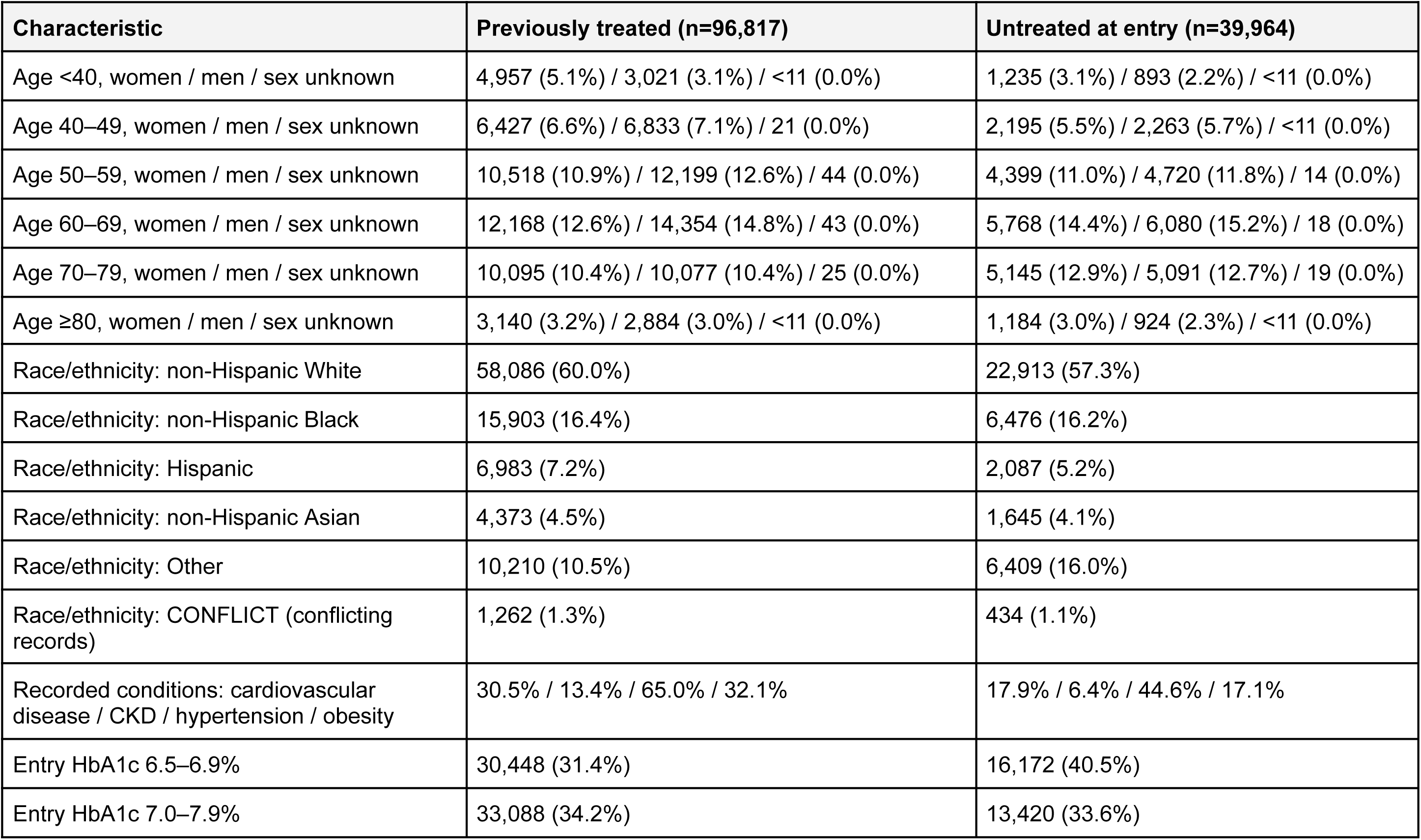
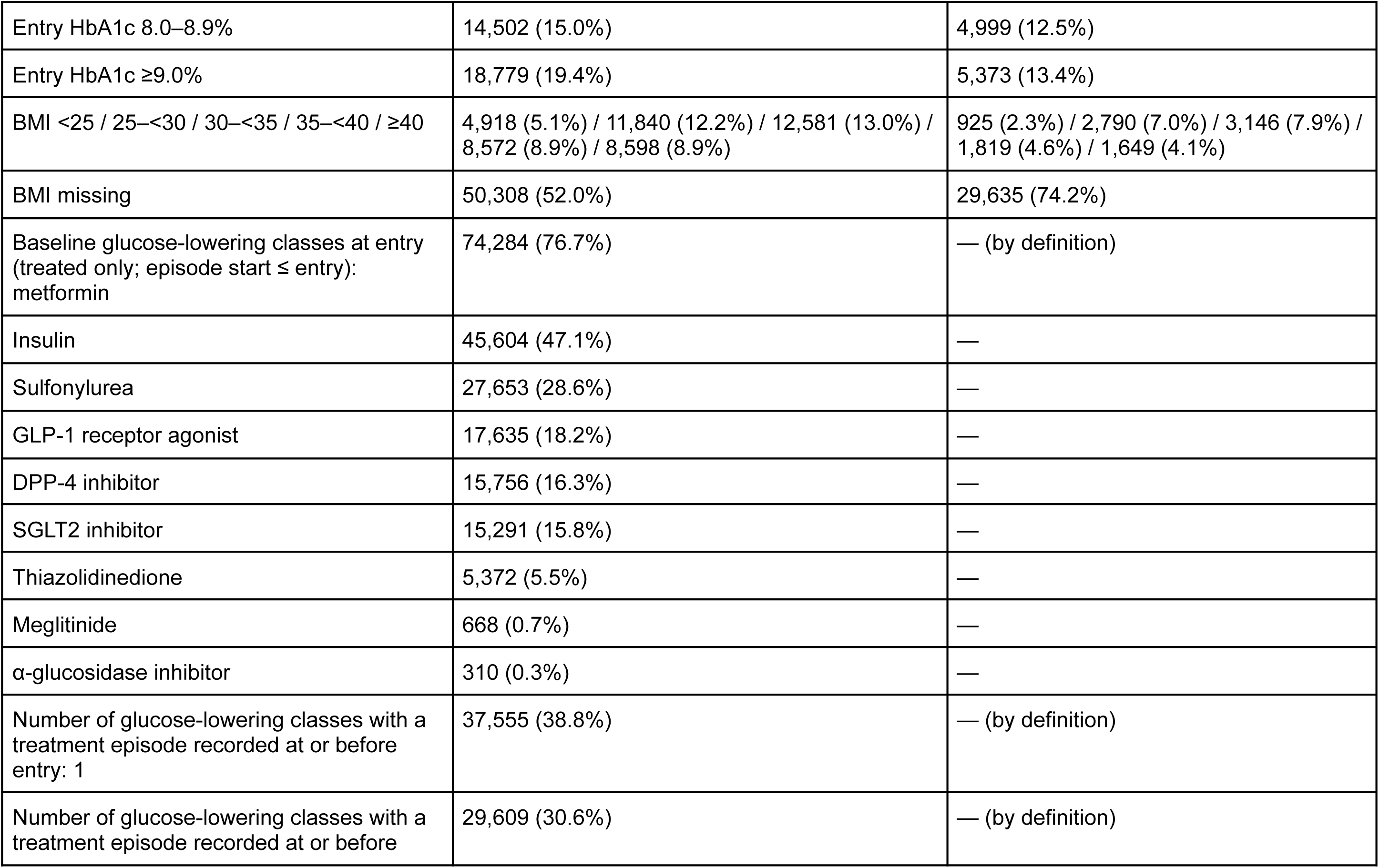
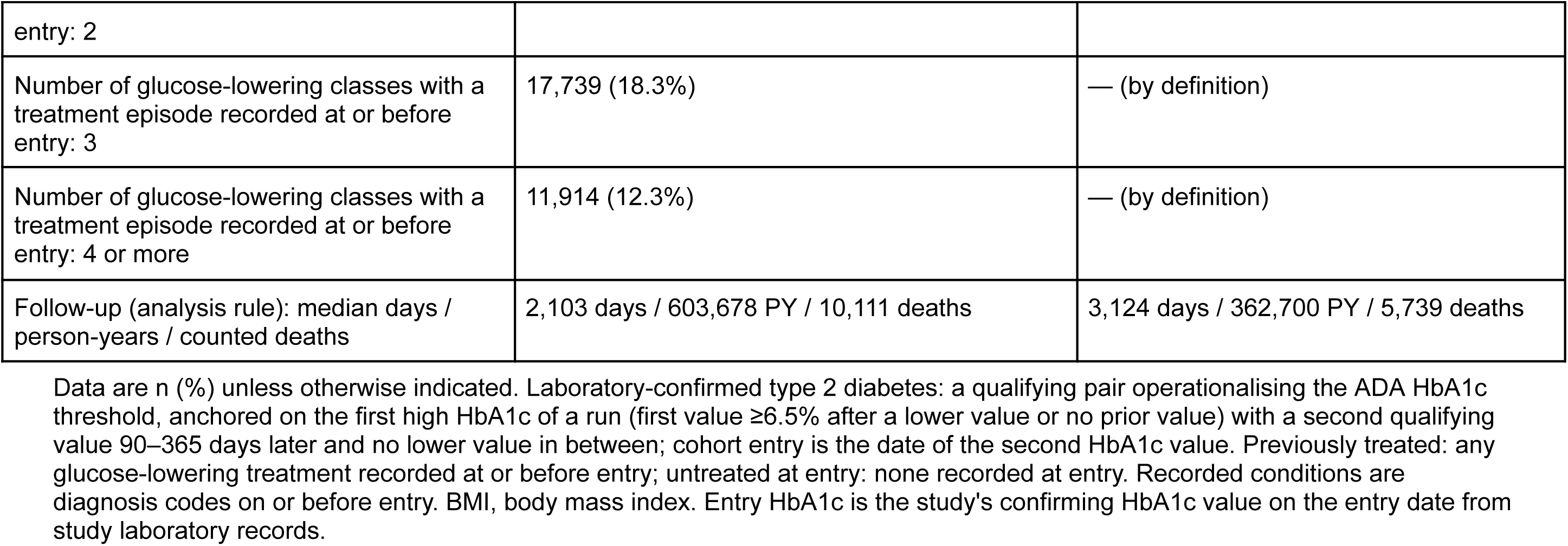
Characteristics at cohort entry of 136,781 adults with laboratory-confirmed type 2 diabetes in a linked EHR and claims cohort, by whether glucose-lowering treatment was recorded at or before entry. Denominators: previously treated 96,817; untreated at entry 39,964.

The primary analytic group was the 96,817 adults with glucose-lowering treatment recorded at or before entry (previously treated cohort). An additional 39,964 adults with no treatment recorded at entry (untreated cohort at entry) were analyzed in parallel.

### Medication records

Treatment periods were constructed for each patient and each of nine glucose-lowering drug classes (metformin, insulin, sulfonylureas, meglitinides, thiazolidinediones, dipeptidyl peptidase 4 inhibitors, SGLT2 inhibitors, GLP-1 receptor agonists including dual GIP/GLP-1 receptor agonists, and α-glucosidase inhibitors) from prescription orders and dispensed pharmacy fills (28). Combination products were counted under each drug class. For treatment cessation a day counted as untreated only if no drug covered the previous 90 days and the record showed that treatment had stopped (pharmacy claims available, a stop order in the past year, or no prior treatment with a recent visit). Cessation could therefore be certified by a planned or supply end date without a recorded stop: among previously treated remitters, 1,264 of 4,188 remissions (30.2%) depended on a planned/supply end date for the last pre-remission episode. Otherwise, the day was classed as unknown, so gaps in the record were not read as stopped treatment. After 2022, when claims ended, restarts were identified from EHR prescriptions, and evidence that treatment had stopped came from stop orders or never-treated status.

### Remission, relapse, and restart

Remission was defined as a first HbA1c below 6.5% at least 90 days after all glucose-lowering drugs were stopped, followed by a second HbA1c below 6.5% 90–365 days later, with no glucose-lowering treatment recorded from 90 days before the first HbA1c through the second (Fig. 1). The remission date was the date of the second HbA1c, with no backdating. Treatment outside the medical record could not be observed, so this is remission as documented, not proof that no medication was taken. Glycemic relapse was the first HbA1c of 6.5% or higher after the remission date, whether or not treatment had been restarted. Treatment restart was the first outpatient prescription or dispensed fill of any glucose-lowering drug after the remission date; drugs given during a hospital stay were recorded but did not count as a restart. End of remission was the first of glycemic relapse, treatment restart, or death. A later remission meeting the same rules after any first end of the first remission counted as a second remission, with death as a competing event. Lasting (sustained) remission at 12, 24, and 60 months required at least two HbA1c tests with no gap longer than 455 days and the last within 455 days of the milestone; patients who did not meet these criteria were classified by the first event (glycemic relapse, treatment restart without relapse, or death) or as having too little testing to classify. For each first treatment restart in the previously treated group, we compared the drug classes restarted with those in the last treatment period before remission. We also recorded weight-loss treatment before remission (GLP-1 receptor agonists, other anti-obesity drugs, or bariatric surgery) and, where weights were available, percent weight change before treatment restart. The remission, relapse, and restart phenotypes were not validated against medical records;

### Deaths

Deaths were counted only if they were dated 2017 or later, were corroborated by a second death record within 31 days or carried a high confidence score, and were not followed by a documented procedure, emergency visit, or hospital stay more than 180 days later. Deaths that failed these checks were not counted, and plausible deaths without corroboration ended follow-up without counting as deaths. Death records that did not meet these checks are summarised in Supplementary Table 3; alternative death definitions were not estimated in the current analysis.

### Statistical analysis

We estimated cumulative incidence with death as a competing event (Aalen–Johansen) (29–31): from cohort entry for remission, and from the remission date for glycemic relapse, end of remission by first event, and second remission, at 1, 2, 3, and 5 years, with 95% CIs and censoring on 30 April 2025. The target was the probability that remission was documented in the record. Adjusted analyses estimated 5-year remission by number of glucose-lowering classes with a treatment episode recorded at or before entry (1, 2, 3, or 4 or more) among previously treated adults and by race and ethnicity within each treatment group. We fitted cause-specific discrete-time hazard models (pooled logistic models with 90-day interval-specific baselines) for remission and for death, and used standardisation (G-computation) to average predicted cumulative incidence over the cohort. Covariates were entry HbA1c band, age band, sex, cardiovascular disease, chronic kidney disease, hypertension, obesity, insulin use, and entry period; race and ethnicity models also included baseline number of drug classes. Body mass index was included in the primary adjusted model with a missing-value indicator. Sensitivity analyses omitted body mass index and, as an exploratory analysis, were restricted to adults with recorded body mass index. Standard errors were obtained by the delta method. These adjusted analyses are descriptive and do not estimate causal effects.

Sensitivity analyses varied the medication-record assumptions (Supplementary Table 4), examined remission by race and ethnicity (Supplementary Table 5), summarised restart by treatment-history tier (Supplementary Table 6), and examined remission by entry period (Supplementary Table 9). Sensitivity analyses using alternative death definitions, alternative confirmation rules including both confirming HbA1c values ≤6.2% [32,33], a single HbA1c value, and two values at least 182 days apart [15], and weighting for HbA1c testing intensity were not performed in the current analysis. An initial protocol and statistical analysis plan were recorded, amendments to the medication definition and the death-record checks were dated. Analyses added after the initial lock, including race × treatment, weight audits, and GLP-1 period analyses, are labelled exploratory. Subgroup results are descriptive, cells with fewer than 11 patients were suppressed, and percentages are reported to one decimal place.

## Results

### Cohort

The 96,817 previously treated adults and 39,964 adults untreated at entry were followed for 603,678 person-years (median 2,103 days) in the previously treated group and 362,700 person-years (median 3,124 days) in the untreated group (Table 1). Previously treated adults entered with poorer glycemic control (19.4% vs 13.4% with HbA1c ≥9.0%) and more often had recorded cardiovascular disease (30.5% vs 17.9%), chronic kidney disease (13.4% vs 6.4%), and hypertension (65.0% vs 44.6%) and obesity (32.1% vs 17.1%). Overall in the previously treated cohort, 60.0% were non-Hispanic White, 16.4% non-Hispanic Black, 7.2% Hispanic, 4.5% non-Hispanic Asian, 10.5% other race or ethnicity and 1.3% had conflict/missing data. BMI was missing in 52.0% of previously treated and 74.2% of untreated adults. Most adults already had diabetes when they entered the cohort. Cohort entry therefore marks the first laboratory confirmation of diabetes in the record, not the date of diagnosis.

### Documentation of T2DM remission

Remission was recorded in 4.33% of previously treated adults (4,188/96,817) and 15.57% of those untreated at entry (6,223/39,964). Five-year cumulative incidence of remission was 3.82% (95% CI 3.69–3.93) in previously treated adults and 13.27% (12.90–13.59) in those untreated at entry, a difference of 9.45 percentage points (Table 2 and Fig. 2A). Sensitivity analyses using alternative medication-record assumptions gave 5-year remission estimates of 2.61% and 12.22% with the discontinuation-evidence sensitivity and 3.71% and 13.15% with the prior-year dispensed-claim sensitivity in previously treated and untreated adults, respectively (Supplementary Table 4; primary cohort, N=136,781). Circumstances before the first treatment restart are summarised in Supplementary Table 7, and deaths by 5 years among adults with sufficient follow-up are summarised in Supplementary Table 8 and Supplementary Fig. 1.

**Figure 2.**
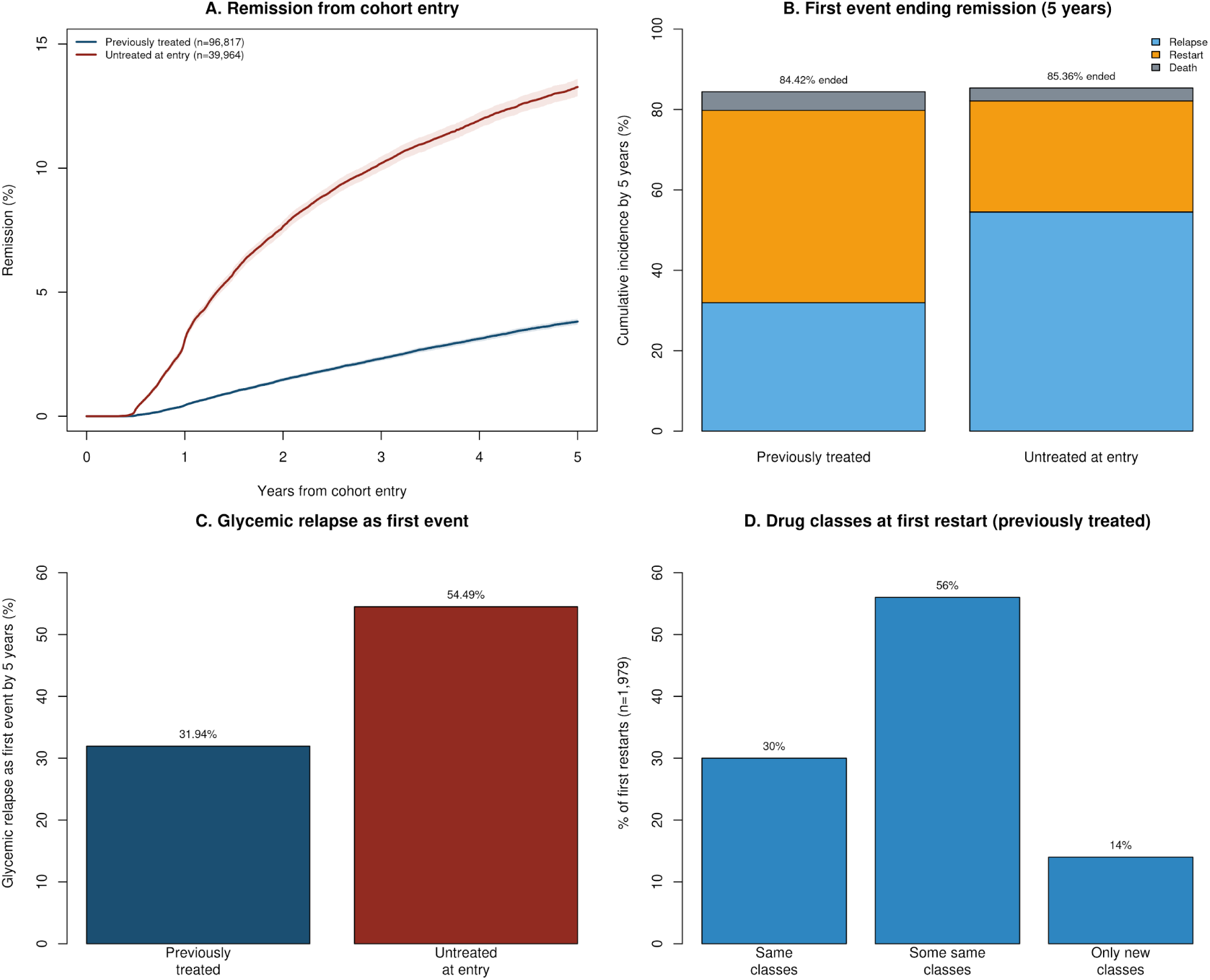
Recorded remission of type 2 diabetes from cohort entry, by treatment history at cohort entry (previously treated, n=96,817; untreated at entry, n=39,964), A: Cumulative incidence of remission (Aalen–Johansen, death as a competing event; shaded bands are 95% CIs). At 5 years: previously treated 3.82% (95% CI 3.69–3.93), untreated at entry 13.27% (12.90–13.59). B: First event ending remission within 5 years of the remission date (glycemic relapse, treatment restart, or death); totals are the cumulative incidence of any end of remission (previously treated 84.42%; untreated at entry 85.36%). C: Glycemic relapse as the first event ending remission by 5 years (previously treated 31.94%; untreated at entry 54.49%). D: Drug classes at the first treatment restart compared with the last treatment period before remission in previously treated adults (n=1,928 restarts with a treatment episode starting on the restart date). Correction: the percentages in the panel D image (30.0% same, 56.0% partial, 14.0% only new; 86.0% any reuse) were calculated against classes ever used before remission; on the stated last-treatment-period comparison the values are 41.9% same, 43.0% partial, and 15.1% only new (84.9% any reuse).

**Table 2.** Cumulative incidence of recorded remission from cohort entry, and of first-event end of remission and second remission from the remission date, among adults with laboratory-confirmed type 2 diabetes, by treatment history at entry. Primary analysis cohort (N=136,781). Sensitivity analyses under alternative definitions are in Supplementary Table 2.

| Outcome | Previously treated(n=96,817) | Untreated at entry (n=39,964) |
| --- | --- | --- |
| Remission ever recorded, % (n/N) | 4.33 (4,188/96,817) | 15.57 (6,223/39,964) |
| Remission, Aalen–Johansen 1 year (52 weeks), % (95% CI) | 0.44 (0.40–0.48) | 3.08 (2.92–3.25) |
| Remission, Aalen–Johansen 2 years (104 weeks), % (95% CI) | 1.47 (1.39–1.54) | 7.65 (7.38–7.88) |
| Remission, Aalen–Johansen 3 years (156 weeks), % (95% CI) | 2.33 (2.23–2.42) | 10.19 (9.90–10.47) |
| Remission, Aalen–Johansen 5 years (260 weeks), % (95% CI) | 3.82 (3.69–3.93) | 13.27 (12.90–13.59) |
| Pooled remission, Aalen–Johansen 5 years, % | 6.71 (both groups combined) |  |
| End of remission by first cause, Aalen–Johansen 5 years: relapse / restart / death, % | 31.94 / 47.83 / 4.65 | 54.49 / 27.65 / 3.22 |
| Any end of remission by 5 years (sum of first causes), % | 84.42 | 85.36 |
| Second remission, Aalen–Johansen 5 years, % | 19.53 | 24.10 |
| First remission episode, Kaplan–Meier median days (IQR) | 226 (93–855) | 371 (166–1,027) |
| Median among episodes observed to end, days (IQR) | 159 (71–362) | 265 (129–565) |

Remission: two HbA1c values <6.5%, 90–365 days apart, the first at least 90 days after all glucose-lowering drugs were stopped, with no drug recorded between them; the remission date is the second value. Glycemic relapse: first HbA1c ≥6.5% after the remission date, whether or not treatment had been restarted. Treatment restart: first outpatient prescription or dispensed fill of any glucose-lowering drug. End of remission: first of relapse, restart, or counted death. Cumulative incidence was estimated with death as a competing event (Aalen–Johansen); 95% CIs for entry remission are bootstrap intervals. Duration medians are Kaplan–Meier estimates (first end = relapse, restart or counted death; censoring at the ledger censor date or 30 April 2025); 37.9% (previously treated) and 23.1% (untreated) of episodes had ended by 152 days (5 months). Medians among episodes that had ended are shown separately and describe only episodes that had ended. Same-day relapse and restart: relapse counted first.

Remission differed modestly by race and ethnicity. At 5 years, cumulative incidence was lowest in Hispanic adults in both treatment groups (3.14% in previously treated and 10.27% in untreated adults, compared with 3.80% and 13.64% in non-Hispanic White adults; Supplementary Table 5). Estimates in non-Hispanic Black adults were 3.56% and 12.09%, in non-Hispanic Asian adults 3.80% and 12.35%, and in other adults 4.78% and 14.46%, respectively. Remission was also more common at lower entry HbA1c: 5-year cumulative incidence was 6.89% for entry HbA1c 6.5–6.9% and 1.47% for ≥9.0% in previously treated adults, and 22.39% and 3.23%, respectively, in adults untreated at entry, declining monotonically across entry HbA1c bands (Supplementary Table 9). Estimates varied by entry period: in previously treated adults, 5-year incidence was 3.84% for entry in 2015 or earlier, 4.17% for 2016–2019, and 2.89% for 2020–2024; in untreated adults it was 11.98%, 14.24%, and 14.64%, respectively (Supplementary Table 9 and Supplementary Fig. 2).

After adjustment for entry HbA1c, age, sex, cardiovascular disease, chronic kidney disease, hypertension, obesity, insulin use, entry period, and body mass index (missing-value indicator), 5-year remission among previously treated adults fell with the number of glucose-lowering classes with a treatment episode recorded at or before entry: 4.65% with 1 such class (n=37,555), 3.62% with 2 classes (n=29,609), 2.69% with 3 classes (n=17,739), and 2.02% with 4 or more classes (n=11,914), compared with unadjusted estimates of 5.29%, 3.56%, 2.43%, and 1.68%, respectively (Supplementary Table 14). Adjusted race and ethnicity differences were small in previously treated adults in the primary analysis and, among adults untreated at entry, lower estimates for Hispanic and non-Hispanic Asian adults were not confirmed in the exploratory complete-case sensitivity analysis (Supplementary Table 14); these comparisons are suggestive. Medication burden (class and/or number of drugs prescribed) at entry also differed from burden immediately before withdrawal among remitters (Supplementary Table 15). Among all 10,411 remitters, 36.7% changed medication burden (30.4% increased and 6.3% decreased); median burden rose from 0 to 1 class, and insulin use rose from 13.5% to 25.0%. Among 4,188 previously treated remitters, 43.5% changed burden and insulin use rose from 33.5% to 42.4%. When insulin was restricted to outpatient sources (outpatient orders and pharmacy dispensed claims, excluding hospital-administered doses), insulin use among all remitters rose from 6.8% at entry to 10.8% immediately before withdrawal (previously treated: 16.8% to 19.9%; untreated at entry: 0.0% to 4.7%), compared with 13.5% to 25.0% using all sources; 56.0% of pre-withdrawal insulin users had only hospital-administered insulin records, so the rise was smaller when hospital doses were excluded (Supplementary Table 15). Among 6,223 adults untreated at entry, 1,998 (32.1%) started treatment after entry and before remission (Supplementary Table 13).

Remission required two HbA1c values below 6.5% recorded 90–365 days apart with no treatment recorded between them (Fig. 1). Sensitivity analyses that varied the medication-record assumptions gave lower 5-year estimates than the primary analysis (Supplementary Table 4). Alternative confirmation rules, including a single HbA1c value and values at least 182 days apart, were not estimated in the current analysis.

Most remissions ended within 5 years. Among previously treated adults, 84.42% were no longer in remission 5 years after the remission date (Table 2 and Fig. 2B). The first event that ended remission was a treatment restart in 47.83%, a glycemic relapse in 31.94%, and death in 4.65%. Under the stricter recorded-discontinuation definition (Supplementary Table 4), restart remained the commonest first event, but the restart/relapse ratio was about 1.2 (restart 42.51%, relapse 34.66%, death 5.46%; ratio 1.23) rather than about 1.5 in the primary analysis. Glycemic relapse as the first event ending remission is shown in Fig. 2C. In adults untreated at entry, the pattern was reversed: relapse came first in 54.49%, treatment restart in 27.65%, and death in 3.22%. Remission was also generally brief. Median Kaplan–Meier duration of the first remission episode was 226 days (IQR 93–855) in previously treated adults and 371 days (166–1,027) in untreated adults; 37.9% (treated) and 23.1% (untreated) of episodes had ended by 5 months (152 days). Among episodes that had ended, median duration was 159 days (71–362) and 265 days (129–565). A second remission followed within 5 years in 19.53% and 24.10%, respectively. Among remitters younger than 85 years with sufficient follow-up, lasting remission at 5 years was documented in 1.81% of previously treated and 3.21% of untreated adults.

### Characterization of Treatment Restarts

Most treatment restarts resumed drugs that the patient was taking before the documented remission. Of 1,928 first treatment restarts in previously treated remitters with a treatment episode starting on the restart date, 84.9% included at least one drug class from the last treatment period before remission: 41.9% (808/1,928) restarted exactly the same classes and 43.0% (829/1,928) some of the same classes, while 15.1% (291/1,928) started only new classes (Fig. 2D; Supplementary Table 10). Metformin alone accounted for 43.9% of restarts; 20.7% included a GLP-1 receptor agonist and 9.2% an SGLT2 inhibitor (Supplementary Table 10). Among restarts with a measured serum glucose between the confirmation date and the restart, the last value was 126 mg/dL or higher in 31.7% of previously treated and 33.2% of untreated remitters. In time-matched comparators the last value was 126 mg/dL or higher in 30.0% in both groups (risk-set sampled; fixed 365-day window 30.3% treated and 30.9% untreated), so a last glucose of 126 mg/dL or higher was slightly more often before restarts than in time-matched comparators (previously treated: 31.7% [92/290] vs 30.0%, +1.7 percentage points; untreated at entry: 33.2% [97/292] vs 30.0%, +3.2 percentage points); the difference was modest (Supplementary Table 7; exploratory, descriptive). Drugs given during a hospital stay were recorded but did not count as a treatment restart (Supplementary Table 2).

#### Weight Change and Treatment Pathways

Paired weights before and after the remission confirmation date were available for 51.9% of previously treated and 50.0% of untreated remitters; weight change in pounds is tabulated in Supplementary Table 11. Prior GLP-1 receptor agonist use was recorded in 372 of 4,188 previously treated remitters (8.9%), including 191 whose last GLP-1 treatment period ended before remission (Supplementary Table 12).

#### Deaths and Calendar Period

Of 20,531 death records in the cohort, 15,850 met the quality checks and were counted; 1,280 were dated before 2017, 561 were followed by a later hospital stay or procedure, and 2,840 lacked corroboration (Supplementary Table 3). Counted deaths were 10,111 in previously treated and 5,739 in untreated adults (Table 1). Among adults with at least 5 years of potential follow-up, 7.07% of previously treated and 3.79% of untreated adults had died by 5 years (Supplementary Table 8). Alternative death definitions were not estimated in the current analysis. Because dates were shifted per patient, calendar periods are approximate. Prior GLP-1 receptor agonist use among previously treated remitters was 3.5% for entry in 2015 or earlier, 9.5% for 2016–2019, and 19.5% for 2020–2024 (Supplementary Table 12).

## Conclusions

Documented remission of type 2 diabetes in routine care is uncommon and brief, and it usually ends because treatment is restarted, not because HbA1c rises. In this cohort of 96,817 previously treated adults, remission was recorded in approximately 1 in 23 patients (4.33%), and half of first remission episodes had ended within 8 months (226 days Kaplan–Meier median; 37.9% by 5 months) in previously treated adults. Within 5 years, 84.42% of remitters were no longer in remission by first event, and 1.81% of those with sufficient follow-up still had lasting remission documented. In addition to estimating how often remission is documented in health records, our study shows how it ends. Treatment restart, usually of a drug class the patient had taken before remission, was the first event about 1.5 times as often as a rise in HbA1c (47.83% vs 31.94% by 5 years), and a last serum glucose of 126 mg/dL or higher was slightly more often before restarts than in time-matched comparators while remission was still documented (31.7% vs 30.0% in previously treated adults and 33.2% vs 30.0% in adults untreated at entry). Under a stricter recorded-discontinuation definition, restart remained the commonest first event, but the restart/relapse ratio was about 1.2 (Supplementary Table 4). Previous real-world studies have combined these two events into a single relapse outcome (19).

Separating glycemic relapse from treatment restart provides a more accurate picture of remission in routine care. Had we counted either event as relapse, we would have concluded that 84.42% of remissions in previously treated adults had ended within 5 years and attributed this to glycemic deterioration. Instead, our data show that most remissions ended with a clinical decision to resume treatment, often when HbA1c remained below 6.5%; a last measured serum glucose of 126 mg/dL or higher was slightly more often before restarts than in time-matched comparators. Clinicians act on fasting glucose, self-monitoring results, and glucose trends, none of which are captured by a dichotomous HbA1c endpoint. Metformin, SGLT2 inhibitors, and GLP-1 receptor agonists are also restarted for kidney, cardiovascular, and weight indications (20). In addition, glucose-lowering drugs are frequently held during hospital admissions and resumed at discharge (34), and in older adults, in whom tight glycemic control may cause harm, treatment is often reduced or stopped and later resumed (35). These decisions reflect appropriate clinical care, and each ends remission without a glycemic relapse.

The same distinction applies to glycemic control while patients remain on treatment. Because the ADA consensus definition counts only remission when patients are not taking any glucose-lowering therapy (1), wider use of GLP-1 receptor agonists would not be expected to increase remission rates, and among previously treated adults in this cohort it did not. Counts of remission after stopping a GLP-1 receptor agonist were small in each calendar period, consistent with the weight regain observed after withdrawal of these drugs in clinical trials (21,22). Few patients had documented weight loss of the magnitude seen in trials, where substantial weight reductions drive remission through decreases in liver and pancreatic fat (2,36). Most documented remissions therefore occurred without recorded weight-loss treatment and instead reflected HbA1c values close to the diagnostic threshold, which may explain why they were brief (Supplementary Table 11).

The operational definitions used to identify remission influenced the estimates as much as glucose physiology did. Remission required two HbA1c values below 6.5%, a range in which biological and analytical variation alone can move a patient back across the diagnostic threshold (32,33). Sensitivity analyses that varied the medication-record assumptions gave 5-year remission estimates of 2.61% and 12.22% with the discontinuation-evidence sensitivity and 3.71% and 13.15% with the prior-year dispensed-claim sensitivity, compared with 3.82% and 13.27% in the primary analysis (Supplementary Table 4; primary cohort). These choices are part of the remission phenotype (24). Our findings support recommending that real-world studies of remission report the confirmation rule, the medication sources and evidence used to establish that treatment had stopped, how periods of unknown treatment status and deaths were handled, and glycemic relapse and treatment restart as separate outcomes.

Using the definitions described in this study, our remission estimates are consistent with previous reports. Our 3-year remission incidence of 2.33% in previously treated adults is close to the 2.9% reported across six U.S. integrated health systems (19) (two consecutive HbA1c values below 6.5% at least 90 days apart). Definitions requiring longer confirmation intervals, as used in the English National Diabetes Audit (15), would be expected to give lower values. The 12-month medication-free definitions used at Kaiser Permanente would be expected to yield lower values (11), and the Scottish prevalence estimate includes diet-controlled patients, who are absent from a laboratory-confirmed cohort (13). The NHS programme in England, which used a programme definition different from the definition used in this study, reported remission in 27% (190/710) among the 42% of starters with two HbA1c values at 12 months after intensive weight management (9); without an intervention, 0.44% of our previously treated cohort had remission recorded by 1 year. Clinical trial results should be interpreted with caution as benchmarks for real-world remission estimates. DiRECT and DIADEM-I enrolled adults with recently diagnosed diabetes, withdrew glucose-lowering drugs at the start of a structured weight-loss program, and measured HbA1c at scheduled visits; DiRECT also excluded insulin users (2–5). Look AHEAD likewise delivered an intensive lifestyle intervention with protocol-based follow-up (6). In contrast, most of our cohort had established diabetes at entry, 47.1% of previously treated patients had insulin recorded, no intervention was delivered, and HbA1c was measured only when clinically indicated. Trial estimates therefore describe what can be achieved with intensive, protocol-driven care, whereas ours describe how often remission is documented in routine practice.

In our parallel analysis of adults untreated at entry, documented remission was more than twice as common as in previously treated adults (13.27% vs 3.82% at 5 years), but this difference should not be interpreted as an effect of treatment. Adults untreated at entry had milder hyperglycemia, and 40.5% entered with an HbA1c between 6.5% and 6.9% (Table 1), where small fluctuations can produce two values below the threshold. Remission in this group also rose across entry periods (Supplementary Fig. 2), consistent with more diagnoses made near the diagnostic threshold in recent years. Their medication history was incomplete by design, because the group was defined only by the absence of treatment recorded at entry; 32.1% (1,998/6,223) of those who reached remission had started glucose-lowering treatment after entry (Supplementary Table 13). Differences by race and ethnicity were modest and unadjusted, and they partly reflected differences in the share of patients untreated at entry. These unadjusted differences may also reflect differences in baseline HbA1c, age, and treatment history, and should not be interpreted as independent associations; adjusted analyses would be needed to assess whether differences persist after accounting for those clinical factors.

Among previously treated adults, differences by race and ethnicity were small in the primary adjusted analysis, apart from a higher estimate in the “Other Race” group; in the exploratory complete-case subset, however, a small Black–White difference was present (−0.99 percentage points). Among adults untreated at entry, the Black–White difference was no longer present in the primary analysis (Supplementary Table 14). The number of glucose-lowering classes with a treatment episode recorded at or before entry remained associated with lower remission after adjustment. Greater treatment burden at entry marked more advanced disease and a lower probability of remission, attenuated after adjustment but persistent, with implications for who is likely to remit and for monitoring after treatment withdrawal. This pattern is best read as a marker of disease severity and treatment need at entry. Among remitters, medication burden at entry also understated burden immediately before withdrawal: 36.7% had a different number of drug classes before withdrawal than at entry (30.4% higher and 6.3% lower), the median rose from 0 to 1 class, and insulin use rose from 13.5% to 25.0% (Supplementary Table 15). Restricted to outpatient insulin sources, insulin use rose from 6.8% to 10.8%, so the rise was smaller when hospital doses were excluded; 56.0% of pre-withdrawal insulin users had only hospital-administered insulin records. Entry medication burden alone therefore does not describe the treatment from which remission was achieved, and burden analyses anchored at entry should be interpreted accordingly; this motivated measuring burden in the last treatment period before withdrawal.

The strengths of this study include a large cohort of patients seeking care in over 20 health systems across the United States, with patient residence in all 50 states and the District of Columbia although four states (California, Virginia, North Carolina, and Ohio) accounted for about 60% of records; Separate tracking of glycemia, treatment, and observation; competing-risk methods; and systematic evaluation of the rules used to ascertain remission, including audits of the medication and death records are also strengths.

On the other hand the study also has several limitations. Protégé is a de-identified EHR and claims resource sampled for research, not a population registry, so our estimates may not generalize to all U.S. adults with type 2 diabetes, and care outside the contributing health systems is not observed. Claims lacked enrollment information and ended in 2022, so treatment may have been incompletely captured, and later restarts relied on EHR prescriptions alone. Medication stops and restarts were inferred from records and were not validated against charts. Death records were imperfect, and alternative death definitions, confirmation rules, and testing-intensity weights were not estimated in the current analysis. Counted deaths are screened records, 20.6% have later activity, and the death file ends Q3 2024 so late deaths are under-ascertained. Dates were shifted for de-identification, making calendar periods approximate, and pregnancies may not have been fully excluded. BMI was often missing, subgroup comparisons were unadjusted, and clustering by the health system could not be modeled. Finally, incomplete HbA1c testing would lead us to underestimate remission, whereas incomplete capture of treatment would lead us to overestimate it, so the net direction of bias is uncertain. The adjusted analyses remain subject to residual confounding, because treatment intensity and recorded diagnoses do not fully capture disease severity or reasons for prescribing. Body mass index was missing for many adults; the primary model used a missing-value indicator, and the exploratory complete-case sensitivity analysis did not confirm the lower adjusted estimates for Hispanic and non-Hispanic Asian adults who were untreated at entry. Those subgroup findings should therefore be interpreted cautiously. Medication burden at entry understated burden immediately before withdrawal in more than one third of remitters, so burden analyses anchored at entry should be interpreted accordingly.

In conclusion, documented remission of type 2 diabetes in routine care was uncommon and short-lived. Most remissions ended within months, and they ended more often because treatment was restarted, usually while HbA1c was still below 6.5%, than because HbA1c rose. For clinicians, these findings suggest that remission achieved without a structured weight-loss program is fragile, and that patients who stop glucose-lowering therapy need continued glucose monitoring and a shared plan for when treatment should resume, consistent with the consensus recommendations (1). For researchers and health systems considering remission as an outcome or quality measure, glycemic relapse and treatment restart should be reported as separate outcomes, together with how remission was confirmed and how treatment cessation was established; without these details, real-world estimates cannot be easily compared or be fully interpreted. Future studies should examine whether sustained remission is associated with lower rates of cardiovascular and kidney complications, and which patients and interventions are most likely to make remission last.

## Acknowledgments

The authors thank the Meta data engineering and Digital Health teams for data access and technical support.

## Use of Artificial Intelligence

This study was supported through using an internal Meta scaffold around Muse Spark 1.3 and Wondert which assisted the authors in all the stages of the study. The model was used to draft the analyses, study cohorts, statistical plan, protocols, and during manuscript preparation. The author drafted all the inputs to the model and all output was verified by the lead author.

## Author Contributions

F.L. conceived and designed the study, supervised the analyses, interpreted the data, and wrote the manuscript. J.D. contributed to the statistical analysis plan and data interpretation. D.K. contributed to interpretation of the data and critical revision of the manuscript. C.C. provided study coordination, data access and interpretation and manuscript review. All authors reviewed and edited the manuscript and approved the final version. F.L. had full access to all the data in the study through the governed analysis environment.

## Data Availability

The Protégé data are not publicly available and access is governed by the data owner. The study protocol, statistical analysis plan, code lists and aggregate results are available from the Corresponding author on reasonable request.

## Supplementary Material

Remission of Type 2 Diabetes in Linked Health Records: Uncommon, Short-Lived, and More Often Ended by Treatment Restart Than by Rising HbA1c. Cohort: 136,781 adults with laboratory-confirmed type 2 diabetes (96,817 previously treated; 39,964 untreated at entry). Cells with fewer than 11 patients are shown as <11. Percentages are reported to one or two decimal places as in the source tables.

**Supplementary Table 1.** Code lists and analytic conventions.

| Element | Definition |
| --- | --- |
| HbA1c | Laboratory result records: 39 laboratory procedure codes plus analyte-name match; NGSP %; plausible range 3.0–20.0%; dates 1 January 2000 to 30 April 2025; same-day EHR and claims duplicates removed. |
| Laboratory confirmation | A qualifying pair operationalising the ADA HbA1c threshold: anchored on the first high HbA1c of a run (first value $\geq 6.5\%$ after a lower value or no prior value), with a second qualifying value 90–365 days later and no lower value in between; cohort entry is the date of the second value. Screen-positive 685,303; laboratory-confirmed 180,894; primary cohort after sequential exclusions 136,781. |
| Exclusions (sequential, age first) | Age <18 years (1,917 removed); other diabetes types (19,722); pregnancy (95); dialysis or kidney failure (1,853); active cancer (12,916); transplant (1,105); steroid treatment at first HbA1c (6,505). |
| Glucose-lowering drug classes (9) | Metformin, insulin, sulfonylurea, meglitinide, thiazolidinedione, DPP-4 inhibitor, SGLT2 inhibitor, GLP-1 receptor agonist (including dual GIP/GLP-1 receptor agonists), α-glucosidase inhibitor; combination products counted under each class. |
| Cardiovascular disease | ICD-10-CM I20–I25, I48, I50, I60–I69 (prefix match), on or before entry. |
| Chronic kidney disease | ICD-10-CM N18, on or before entry. |
| Hypertension | ICD-10-CM I10–I13, I15–I16 (I14 excluded), on or before entry. |
| Obesity | ICD-10-CM E66, on or before entry. |
| Remission | Two HbA1c values <6.5%, 90–365 days apart, the first at least 90 days after all glucose-lowering drugs were stopped, with no drug recorded between them; remission date is the second value. |
| Conventions | Default order supply 90 days; records chained within 180 days; stop-order window 365 days; person-years = follow-up days/365.25; last HbA1c carried forward up to 455 days for lasting-remission classification. |

DPP-4, dipeptidyl peptidase 4; GIP, glucose-dependent insulintropic polypeptide; GLP-1, glucagon-like peptide 1; NGSP, National Glycohemoglobin Standardization Program; SGLT2, sodium–glucose cotransporter 2. Full code lists are available from the corresponding author on reasonable request.

**Supplementary Table 1A.**
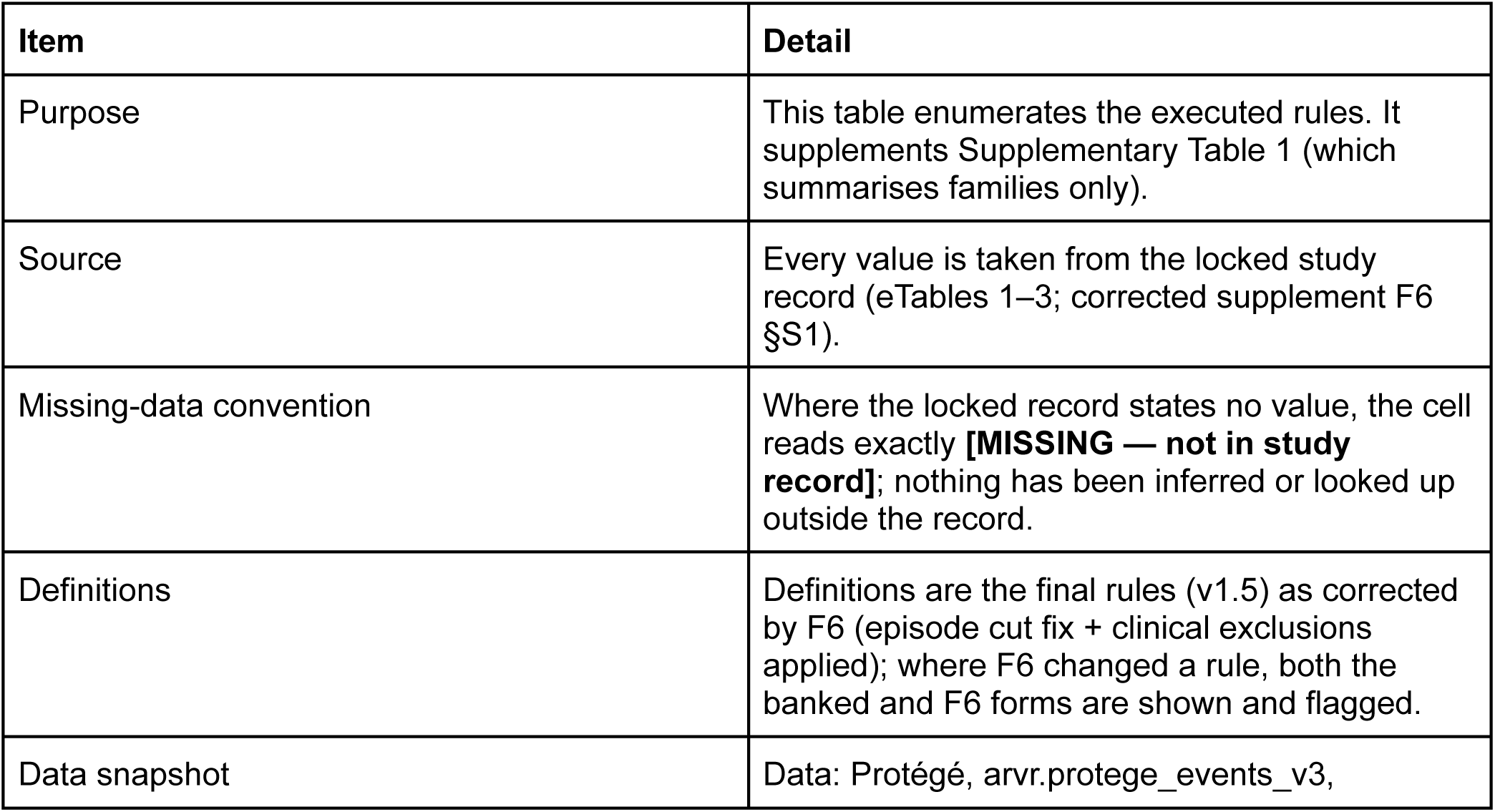
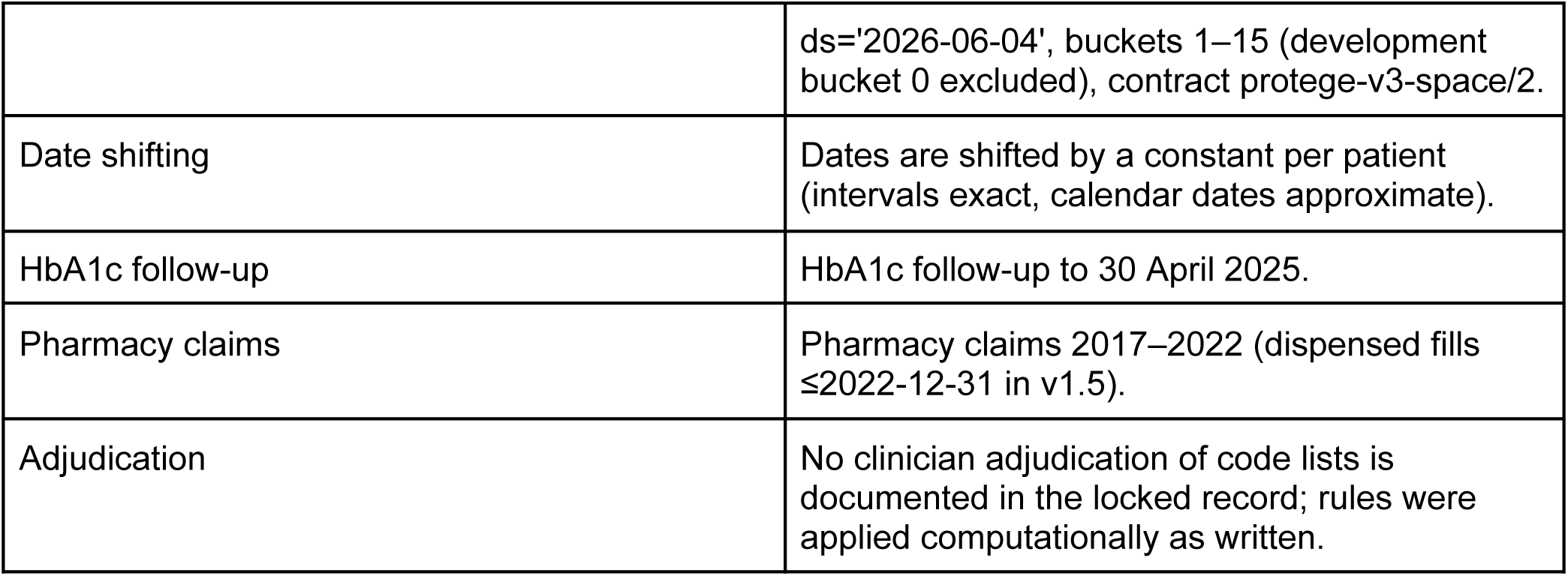
Code lists and algorithms used to define the cohort, exposures, outcomes, and covariates. **Supplementary Table 1 A — scope and provenance**

**Table 1A.1.** HbA1c identity — all 39 laboratory procedure codes (executed)

| # | LABPX code (exact, IN list) | Analyte / unit |
| --- | --- | --- |
| 1 | LAB90 | HbA1c (NGSP %) |
| 2 | LAB100502 | HbA1c (NGSP %) |
| 3 | 2356 | HbA1c (NGSP %) |
| 4 | POC4 | HbA1c (NGSP %) |
| 5 | LAB2172 | HbA1c (NGSP %) |
| 6 | LABP000212 | HbA1c (NGSP %) |
| 7 | A1C | HbA1c (NGSP %) |
| 8 | LAB90HT | HbA1c (NGSP %) |
| 9 | LAB5800 | HbA1c (NGSP %) |
| 10 | LAB684 | HbA1c (NGSP %) |
| 11 | POC83036 | HbA1c (NGSP %) |
| 12 | 999EXTHGBA1 | HbA1c (NGSP %) |
| 13 | LAB12962 | HbA1c (NGSP %) |
| 14 | GENLAB | HbA1c (NGSP %) |
| 15 | LAB90210 | HbA1c (NGSP %) |
| 16 | LAB2740 | HbA1c (NGSP %) |
| 17 | 1231000369 | HbA1c (NGSP %) |
| 18 | LAB10382 | HbA1c (NGSP %) |
| 19 | HXLAB113 | HbA1c (NGSP %) |
| 20 | HXLAB24711 | HbA1c (NGSP %) |
| 21 | POC500002 | HbA1c (NGSP %) |
| 22 | POC81007 | HbA1c (NGSP %) |
| 23 | 83036 | HbA1c (NGSP %) |
| 24 | POC14016 | HbA1c (NGSP %) |
| 25 | LAB10004 | HbA1c (NGSP %) |
| 26 | LAB90211 | HbA1c (NGSP %) |
| 27 | 76546 | HbA1c (NGSP %) |
| 28 | POC124 | HbA1c (NGSP %) |
| 29 | HXLAB4777 | HbA1c (NGSP %) |
| 30 | POC108 | HbA1c (NGSP %) |
| 31 | LAB686 | HbA1c (NGSP %) |
| 32 | LAB2271 | HbA1c (NGSP %) |
| 33 | LABSC00021 | HbA1c (NGSP %) |
| 34 | POC122 | HbA1c (NGSP %) |
| 35 | LAB91004 | HbA1c (NGSP %) |
| 36 | LABC2356 | HbA1c (NGSP %) |
| 37 | A1CRPD | HbA1c (NGSP %) |
| 38 | LAB3862 | HbA1c (NGSP %) |
| 39 | POC264 | HbA1c (NGSP %) |

**Table 1A.2.**
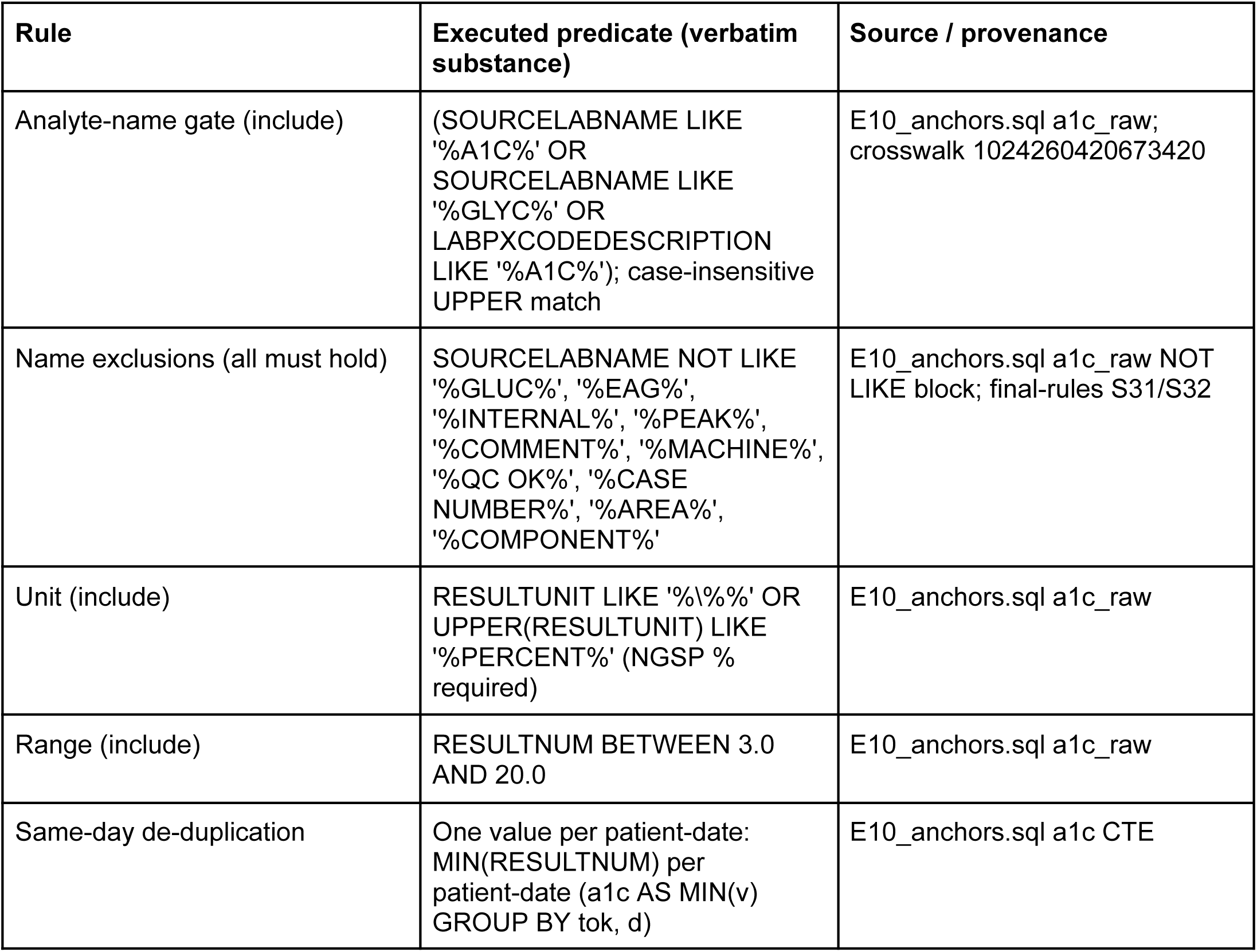
HbA1c analyte-name, unit, range, and de-duplication rules (executed, applied with AND)

**Table 1A.3.**
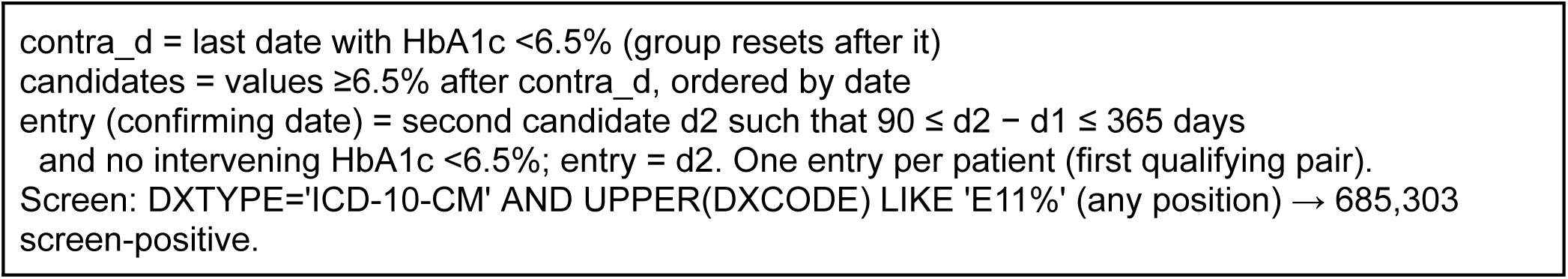
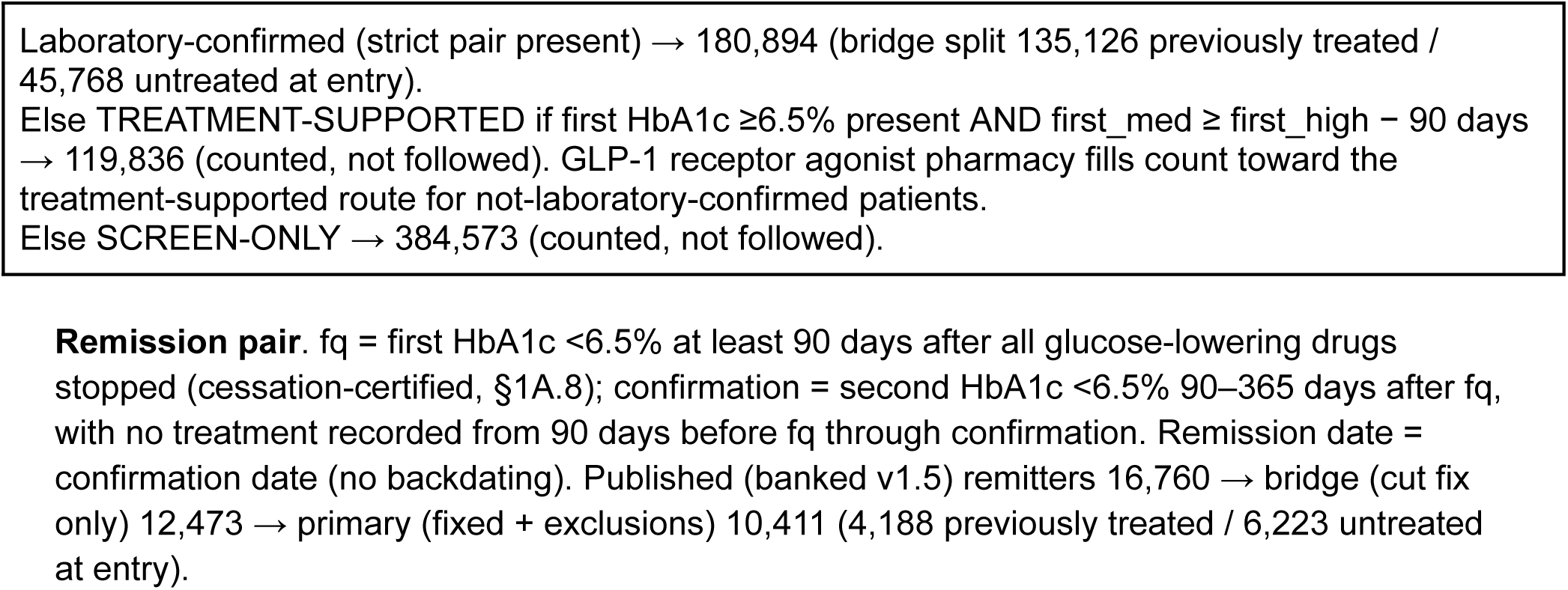
Anchored pairing algorithms (pseudocode, as executed) **Diagnostic (cohort-entry) pair**. Let H(p) be the per-patient-date MIN HbA1c series after

**Table 1A.4.**
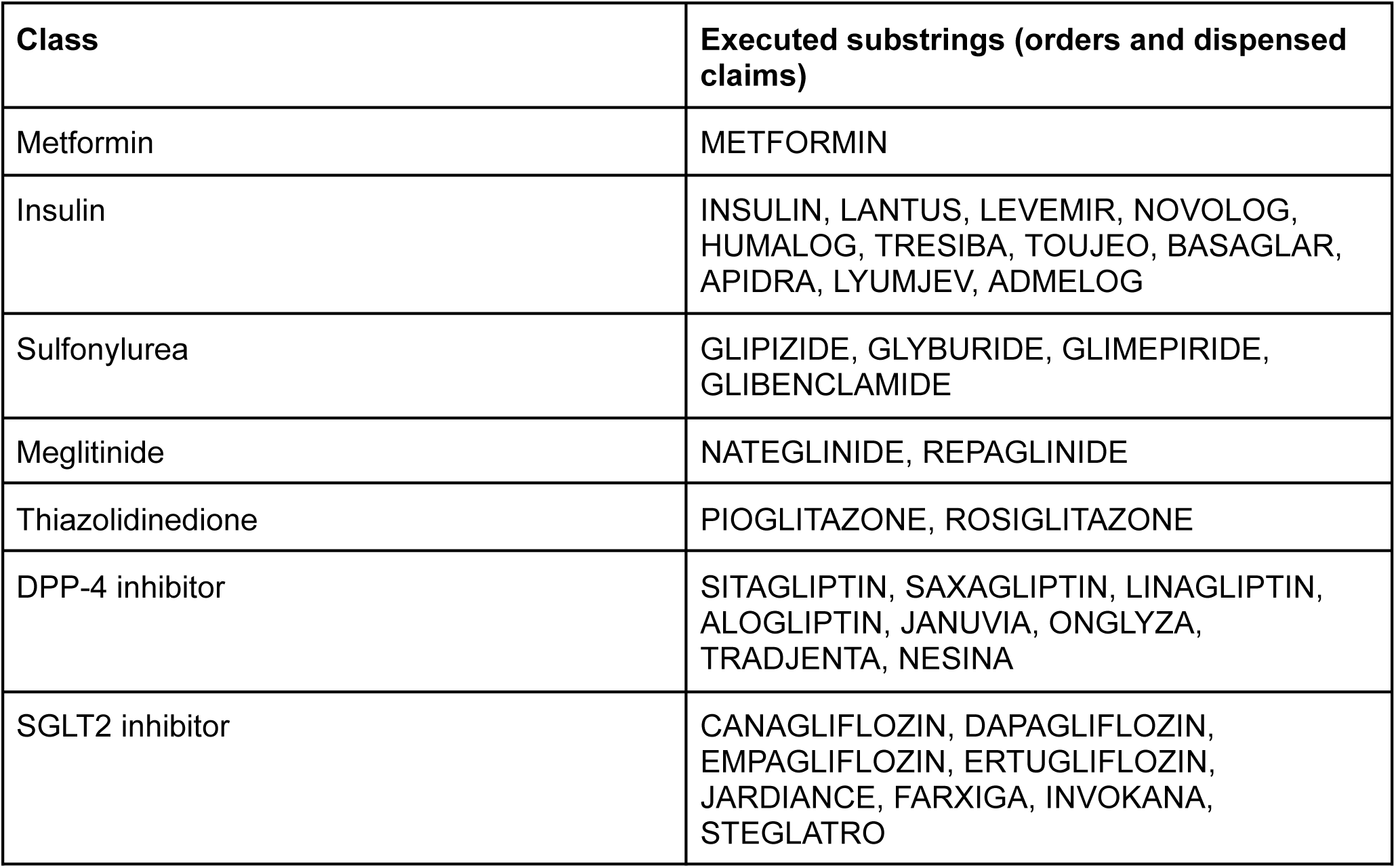
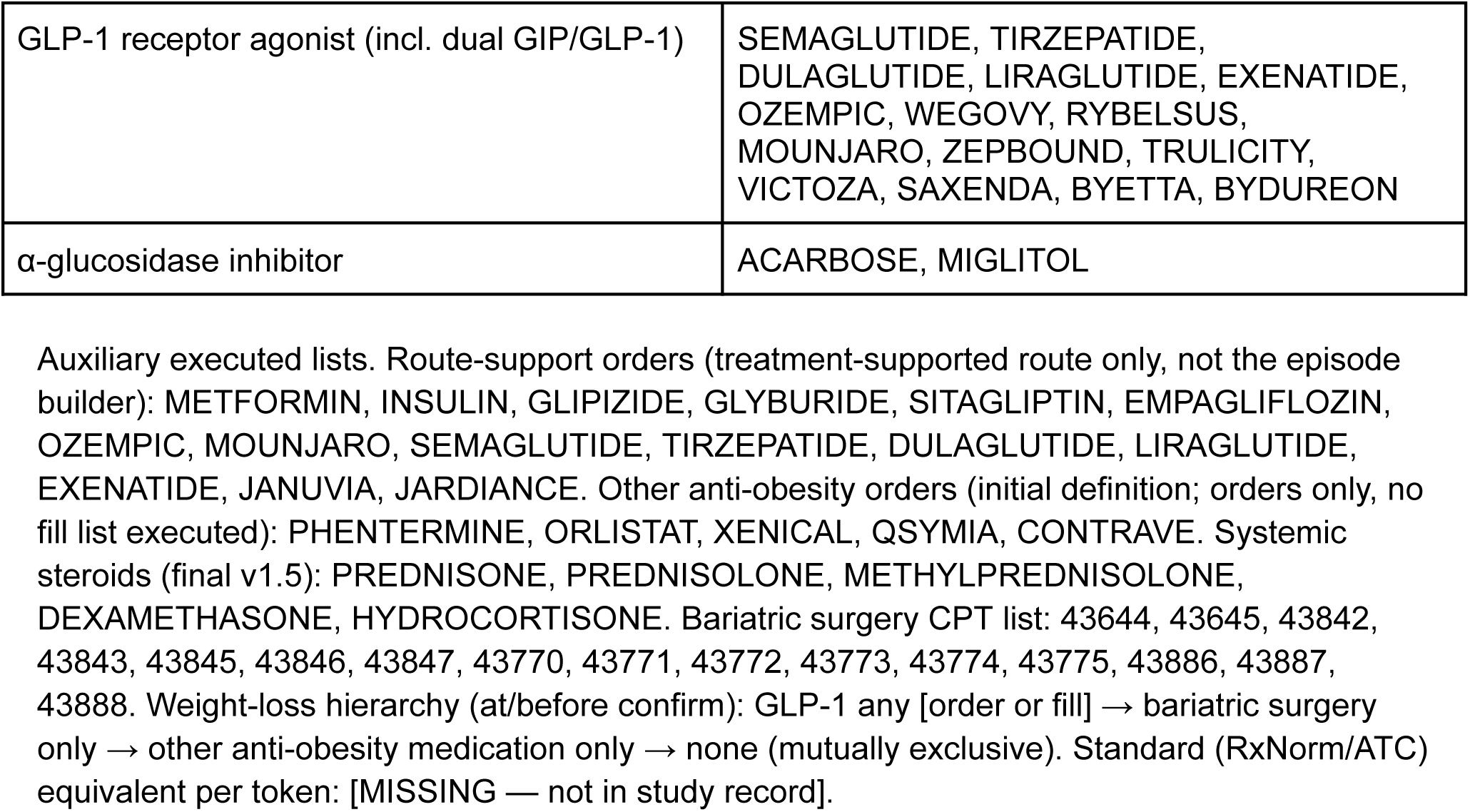
Glucose-lowering drug classes — ingredient (substring) lists as executed. Combination products count under each class they match. Treated at entry = any episode with ep_start ≤ entry (same-day = treated). The executed classifier expands beyond the listed ingredient substrings to brand-name products (for example JANUMET, GLUCOPHAGE);

**Table 1A.5.** Comorbidity ICD-10-CM families (at/before entry, EHR only)

| Flag | Executed prefixes (any position; hai diagnosis, dx_date ≤ entry) | Notes |
| --- | --- | --- |
| Cardiovascular disease | I20%, I21%, I22%, I23%, I24%, I25%, I48%, I50%, I60%, I61%, I62%, I63%, I64%, I65%, I66%, I67%, I68%, I69% | Ischaemic heart disease I20–I25, AF/flutter I48, heart failure I50, cerebrovascular I60–I69 |
| Chronic kidney disease | N18% | N18.1–N18.6, N18.9 and any N18 descendant; labs not used |
| Hypertension | I10%, I11%, I12%, I13%, I15%, I16% (I14% intentionally excluded) | Vitals BP not used |
| Obesity (recorded) | E66% | Does not replace measured BMI |

Presence only (recorded vs absent; no adjudication/severity). ICD-10-CM version: [MISSING — not in study record]. Matched descendant expansion: [MISSING — not in study record].

**Table 1A.6.** Death algorithm (screened death, primary)

| Component | Executed rule |
| --- | --- |
| Counted death (all must hold) | FACT_OF_DEATH dated $\geq 2017$ AND (second death record within 31 days OR confidence score $\geq 94$) AND no procedure / ED / inpatient stay $> 180$ days after death. WA/AK/MT/ID/HI/OR DECEASEDDATE corroboration suppressed. Corroboration means a second death record 0-31 days AFTER the death date (one-sided), not within +/-31 days. |
| Dated before 2017 | Not counted; follow-up continues (bridge 1,618; primary 1,280) |
| Activity-screen fail ($> 180$d activity) | Not counted; follow-up continues (bridge 778; primary 561) |
| Plausible but uncorroborated | Censored at recorded death date, not counted (bridge 3,747; primary 2,840) |
| Totals | Bridge: 27,313 recorded → 21,170 counted. Primary cohort: 20,531 recorded → 15,850 counted (10,111 previously treated / 5,739 untreated); alive at censoring 116,250. Administrative censor 2025-04-30 (shifted). |

**Supplementary Table 2.** Sensitivity analyses: remission under alternative episode and cohort definitions.

| Measure | Primary analysis (N=136,781) | Alternative episode definition, full laboratory-confirmed cohort (N=180,894) | Earlier published banked v1.5 definition (N=180,894) |
| --- | --- | --- | --- |
| Cohort (previously treated / untreated at entry) | primary 136,781 (96,817 / 39,964) | 180,894 (135,126 / 45,768) | 180,894 (135,126 / 45,768) |
| Remitters (previously treated / untreated at entry) | 4,188 / 6,223 | 5,398 / 7,075 | 8,721 / 8,039 |
| Ever recorded, previously treated / untreated at entry, % | 4.33 / 15.57 | 3.99 / 15.46 | 6.45 / 17.56 |

| Measure | Primary analysis<br>(N=136,781) | Alternative episode<br>definition,<br>full laboratory-confirmed<br>cohort (N=180,894) | Earlier published<br>banked v1.5<br>definition<br>(N=180,894) |
| --- | --- | --- | --- |
| 5-year cumulative incidence<br>(Aalen–Johansen), previously<br>treated / untreated at entry, % | 3.82 (95% CI<br>3.69–3.93) / 13.27<br>(12.90–13.59) | 3.65 (3.55–3.78) / 13.34<br>(13.02–13.65) | 5.90 / 14.53 |
| First event by 5 years in previously<br>treated adults: relapse / restart /<br>death, % | 31.94 / 47.83 / 4.65 | 31.94 / 47.36 / 5.01 | 24.4 / 60.8 / 4.0 |
| Median first remission episode,<br>previously treated, days | 226 (Kaplan–Meier) | 159 (among ended<br>episodes) | 108 (among ended<br>episodes) |
| Counted deaths in cohort | 15,850 | 21,170 | 21,170 |

The alternative episode definition applies the same stopping rule with an extended episode end; the primary analysis additionally applies the sequential exclusions and the age 18 years or older requirement. Earlier published estimates are shown only in this sensitivity table to quantify the effect of the definition and cohort choices. For the primary analysis, 226 days is the Kaplan–Meier median; 159 days is the median among episodes that had ended. In the other columns, 159 and 108 days are likewise medians among episodes that had ended, not Kaplan–Meier medians.

**Supplementary Table 3.** Quality checks applied to death records.

| Classification | n | Handling |
| --- | --- | --- |
| Recorded deaths in the cohort (fact-of-death records) | 20,531 | Starting records. |
| Counted deaths | 15,850 | Dated 2017 or later, corroborated by a second record within 31 days or a high confidence score, and not followed by a procedure, emergency visit, or hospital stay more than 180 days later. Previously treated 10,111; untreated at entry 5,739. |
| Dated before 2017 | 1,280 | Not counted as deaths; follow-up ended at the last clinical activity. |
| Followed by clinical activity more than 180 days later | 561 | Not counted as deaths; follow-up ended at the last clinical activity. |
| Plausible but not corroborated | 2,840 | Follow-up ended at the recorded date without counting as a death. |
| Alive at censoring | 116,250 | Administrative censor 30 April 2025 (shifted). |

**Supplementary Table 4.** Remission under alternative medication-record assumptions (sensitivity analyses, primary cohort N=136,781)

| Analysis | Previously treated: remitters; ever, %; 5-year cumulative incidence, % | Untreated at entry: remitters; ever, %; 5-year cumulative incidence, % |
| --- | --- | --- |
| Primary analysis | 4,188; 4.33; 3.82 (95% CI 3.69–3.93) | 6,223; 15.57; 13.27 (12.90–13.59) |
| Discontinuation-evidence sensitivity (stricter recorded-discontinuation cessation ledger) | 2,923; 3.02; 2.61 (95% CI 2.50–2.72) | 5,615; 14.05; 12.22 (11.89–12.55) |
| Prior-year dispensed-claim sensitivity | 4,121; 4.26; 3.71 (3.58–3.84) | 6,183; 15.47; 13.15 (12.81–13.49) |
| Pharmacy-fill sensitivity using EHR fills | Not estimated | Not estimated |

The pharmacy-fill sensitivity using EHR fills was not estimated because the required dispensing fields were not available for analysis. All rows use the primary cohort (N=136,781: 96,817 previously treated; 39,964 untreated at entry). The discontinuation-evidence row is the stricter recorded-discontinuation cessation ledger restricted to the primary cohort; it is not a dispensed-only analysis. Differences in 5-year Aalen–Johansen incidence vs primary were –1.21 (treated) and –1.05 (untreated) percentage points for the discontinuation-evidence sensitivity and –0.11 and –0.12 for the prior-year dispensed-claim sensitivity. Under the stricter recorded-discontinuation (discontinuation-evidence) ledger, the first event by 5 years among previously treated remitters was glycemic relapse in 34.66%, treatment restart in 42.51%, and death in 5.46% (restart/relapse ratio 1.23, compared with 1.50 in the primary analysis); restart remained the commonest first event, ratio 1.2 r

**Supplementary Table 5.** Remission in population subgroups and restricted cohorts.

| Race and ethnicity (modal record) | Previously treated: n; 5-year cumulative incidence, % | Untreated at entry: n; 5-year cumulative incidence, % |
| --- | --- | --- |
| Non-Hispanic White | 58,086; 3.80 | 22,913; 13.64 |
| Non-Hispanic Black | 15,903; 3.56 | 6,476; 12.09 |
| Hispanic | 6,983; 3.14 | 2,087; 10.27 |
| Non-Hispanic Asian | 4,373; 3.80 | 1,645; 12.35 |
| Other | 10,210; 4.78 | 6,409; 14.46 |
| Conflicting records | 1,262; 4.27 | 434; 11.49 |

Race and ethnicity are the modal recorded values; conflicting records are retained as a separate group. Race and ethnicity comparisons were descriptive and should not be interpreted as causal effects. Differences in treatment history, missing covariate information, and ascertainment may affect these comparisons. Exploratory adjusted estimates varied across model and missing-data specifications and did not establish a consistent pattern of differences across groups. Other restricted cohorts were not estimated in the current analysis.

**Supplementary Table 6.** Sensitivity of remission to unknown treatment time and restart by treatment-history tier.

| Tier (previously treated remitters) | Remitters, n | Restart by 1 year, % (n/N observed) | Restart by 5 years, % (n/N observed) | Restart by 1 year with no unknown days, % (n/N) | Median days to restart |
| --- | --- | --- | --- | --- | --- |
| Tier E | 2,211 | 54.17 (1,105/2,040) | 80.30 (970/1,208) | 54.01 (808/1,496) | 96 |
| Tier S | 1,977 | 36.32 (710/1,955) | 64.16 (324/505) | 37.32 (584/1,565) | 116 |

Tier definitions: Tier E = episode-anchored evidence; Tier S = stop-order evidence. Tiers classify the evidence that treatment had stopped before remission. Denominators require full follow-up to the horizon (remission date plus 365 or 1,825 days on or before 30 April 2025). The full unknown-time sensitivity was not estimated in the current analysis; the no-unknown-days columns show the restricted estimates.

**Supplementary Table 7.**
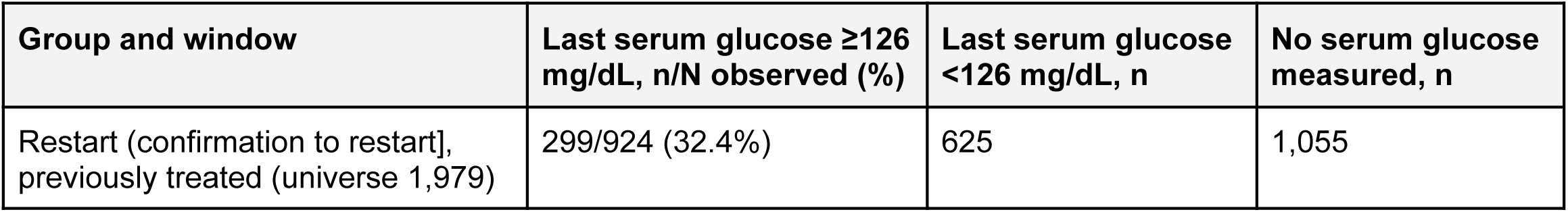
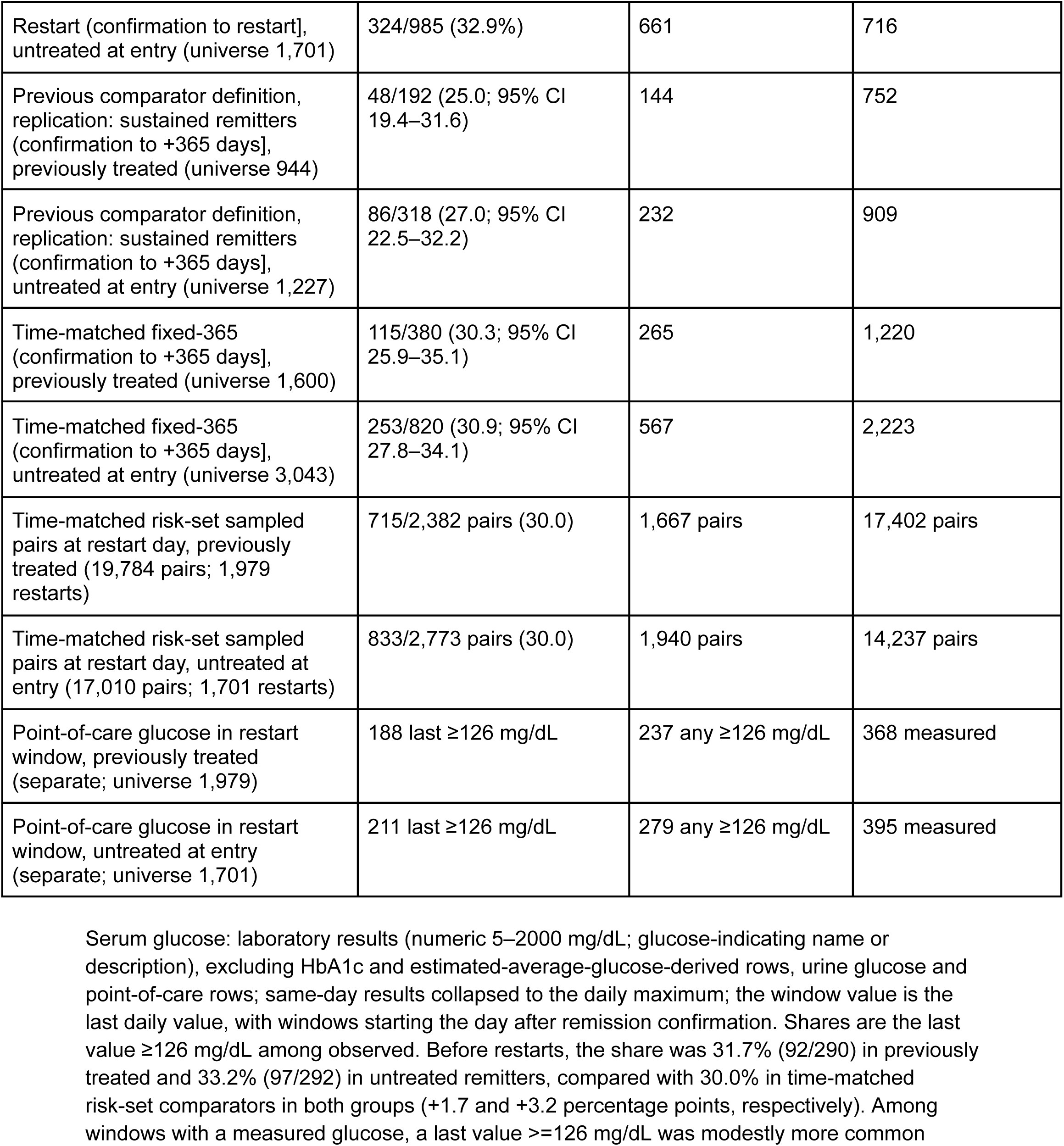
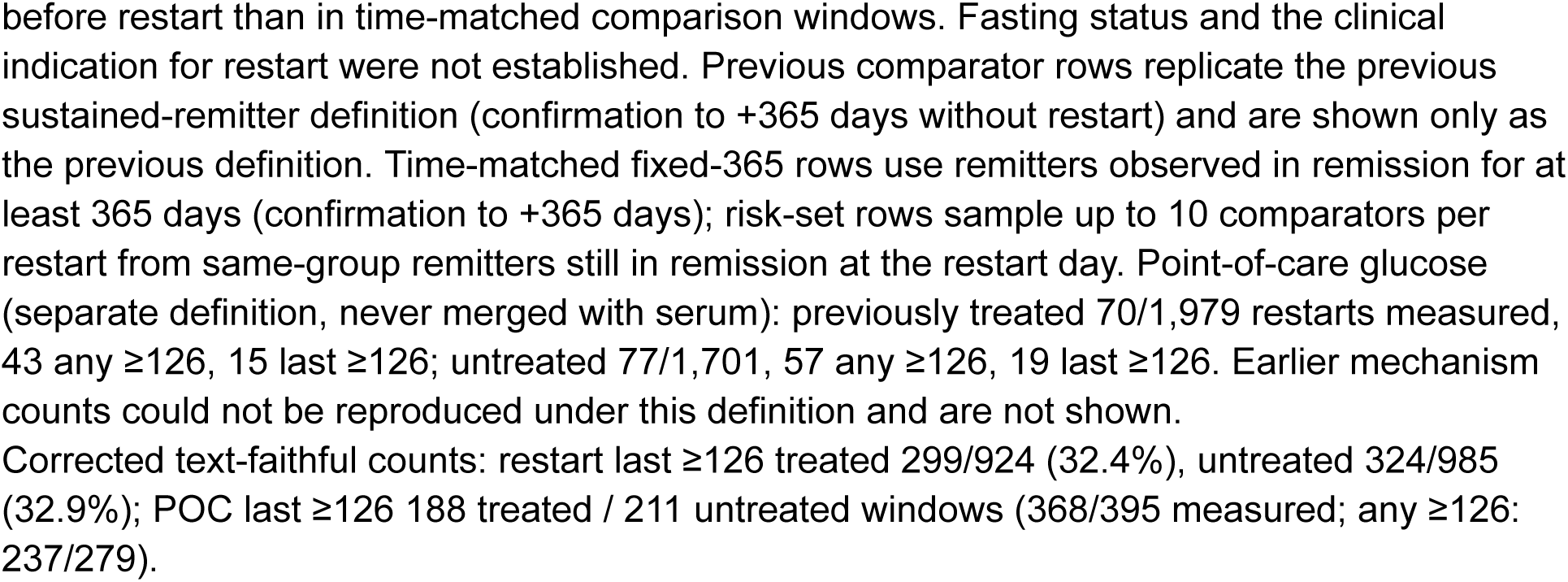
Circumstances before the first treatment restart.

**Supplementary Table 8.** Glycemic and treatment states after cohort entry and lasting remission.

| Measure | Previously treated | Untreated at entry |
| --- | --- | --- |
| Adults with at least 5 years of potential follow-up, n | 61,237 | 32,783 |
| Dead by 5 years, n (%) | 4,332 (7.07) | 1,243 (3.79) |
| Lasting remission at 1 year among remitters younger than 85 years, % (n/N observed) | 25.41 (1,014/3,991) | 34.82 (2,098/6,026) |
| Lasting remission at 2 years, % (n/N observed) | 13.54 (472/3,486) | 18.60 (1,034/5,559) |
| Lasting remission at 5 years, % (n/N observed) | 1.81 (31/1,713) | 3.21 (121/3,767) |

Lasting remission required at least two HbA1c tests with no gap longer than 455 days and the last test within 455 days of the milestone, with no HbA1c $\geq 6.5\%$ and no treatment restart. Full glycemic and treatment state occupancy at landmarks was not estimated in the current analysis; only deaths and lasting remission are reported.

**Supplementary Table 9.** Remission by entry period, age and sex, entry HbA1c, and BMI.

| Entry period | Previously treated: 5-year cumulative incidence, % | Untreated at entry: 5-year cumulative incidence, % |
| --- | --- | --- |
| 2015 or earlier | 3.84 | 11.98 |
| 2016–2019 | 4.17 | 14.24 |
| 2020–2024 | 2.89 | 14.64 |
| Entry HbA1c 6.5–6.9% | 6.89 | 22.39 |
| Entry HbA1c 7.0–7.9% | 3.35 | 9.76 |
| Entry HbA1c 8.0–8.9% | 1.81 | 5.20 |
| Entry HbA1c ≥9.0% | 1.47 | 3.23 |

**Supplementary Table 10.** First event after remission and drug classes at the first treatment restart, by treatment history at entry.

| Measure | Previously treated | Untreated at entry |
| --- | --- | --- |
| Remitters, N | 4,188 | 6,223 |
| First event by 5 years (Aalen–Johansen): relapse / restart / death, % | 31.94 / 47.83 / 4.65 | 54.49 / 27.65 / 3.22 |
| First exit over all follow-up (counts): restart, n (%) | 1,979 (47.3) | 1,701 (27.3) |
| Recurrence (HbA1c ≥6.5%), n (%) | 1,115 (26.6) | 3,086 (49.6) |
| Death, n (%) | 150 (3.6) | 209 (3.4) |
| No exit recorded (sustained in record), n (%) | 944 (22.5) | 1,227 (19.7) |
| Second remission by 5 years (Aalen–Johansen), % | 19.53 | 24.10 |
| Restarts with drug-class comparison, n | 1,928 | 1,701 (753 with an earlier treatment period) |
| Exactly the same classes, % (n) | 41.9 (808) | 37.2 (280/753 with earlier period) |
| Some of the same classes (partial overlap), n | 829 | 364 |
| Only new classes, n | 291 | 109 |
| No earlier treatment period, n | — | 948 |
| Any earlier class reused, % | 84.9 | 85.5 (of those with an earlier period) |
| Metformin only at restart, n | 869 | 776 |
| Restart including a GLP-1 receptor agonist, n | 417 | 280 |
| Restart including an SGLT2 inhibitor, n | 182 | 134 |

**Supplementary Table 11.**
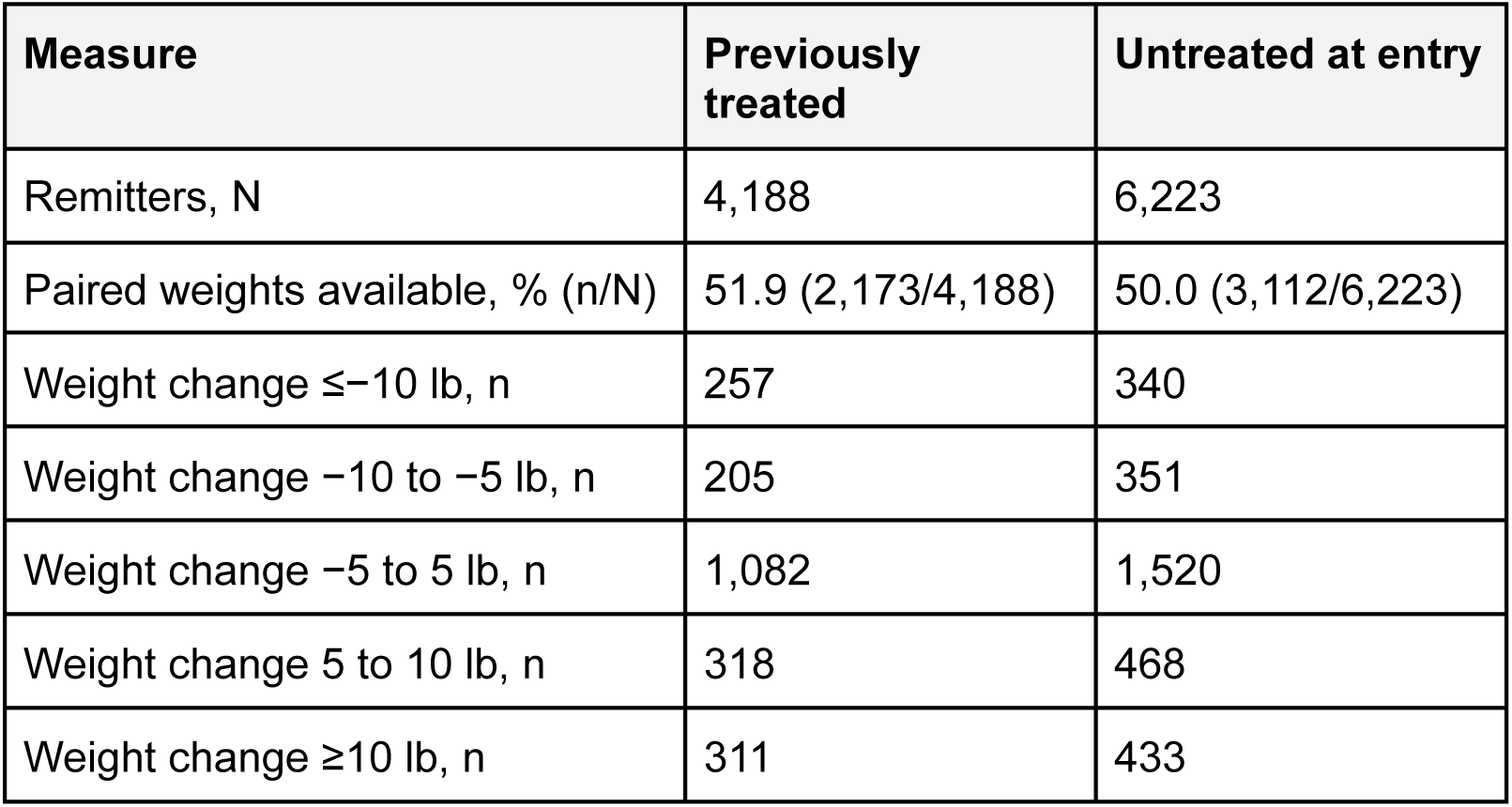
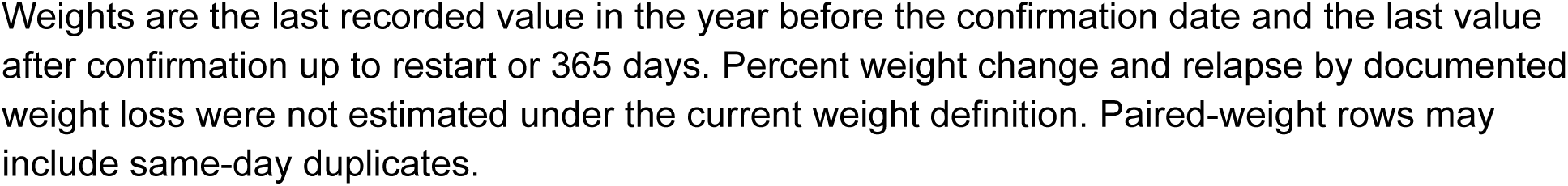
Weight-loss treatment before remission and weight change.

**Supplementary Table 12.** GLP-1 receptor agonist use and remission after GLP-1 receptor agonist cessation, by entry period.

| **Entry period and group** | **Cohort, n** | **Prior GLP-1 use in cohort, n** | **Remitters, n** | **Remitters with prior GLP-1 use, n (%)** | **Remitters whose last GLP-1 period ended before remission, n** |
| --- | --- | --- | --- | --- | --- |
| 2015 or earlier, previously treated | 25,216 | 1,507 | 1,757 | 62 (3.5) | 29 |
| 2015 or earlier, untreated at entry | 19,932 | 0 | 3,368 | 0 | 0 |
| 2016–2019, previously treated | 32,664 | 4,324 | 1,636 | 155 (9.5) | 71 |
| 2016–2019, untreated at entry | 11,968 | 0 | 1,895 | 0 | 0 |
| 2020–2024, previously treated | 38,937 | 11,804 | 795 | 155 (19.5) | 91 |
| 2020–2024, untreated at entry | 8,064 | 0 | 960 | 0 | 0 |

Prior use is a GLP-1 treatment period starting on or before cohort entry. Glycemic control on treatment at 2 years was not estimated in the current analysis. Periods use per-patient shifted dates and are approximate.

**Supplementary Table 13.** Remitters who started treatment before remission vs those untreated through remission.

| Group | Started treatment before remission, n (%) | Untreated through remission, n |
| --- | --- | --- |
| Untreated at entry who reached remission (N=6,223) | 1,998 (32.1) | 4,225 |

Started treatment means any glucose-lowering treatment recorded between cohort entry and the first qualifying HbA1c. Relapse comparisons by this grouping were not estimated in the current analysis.

**Supplementary Table 14.** Adjusted 5-year remission by glucose-lowering classes with a treatment episode recorded at or before entry and race and ethnicity. Adjusted estimates used cause-specific discrete-time hazard models with standardisation (G-computation), adjusting for entry HbA1c band (study rule), age, sex, cardiovascular disease, chronic kidney disease, hypertension, obesity, insulin use, entry period, and body mass index with a missing-value indicator; race and ethnicity models also adjusted for the number of glucose-lowering classes with a treatment episode recorded at or before entry. Sensitivity analyses omitted body mass index and, exploratorily, were restricted to adults with recorded body mass index. The complete-case sensitivity analysis did not confirm the lower adjusted estimates for Hispanic and non-Hispanic Asian adults untreated at entry. In the sensitivity analysis omitting body mass index, adjusted differences for adults untreated at entry were −1.69 percentage points for Hispanic and −1.96 for non-Hispanic Asian adults (vs non-Hispanic White). In the exploratory complete-case analysis restricted to adults with recorded body mass index (a selected subset comprising 26% of adults untreated at entry), adjusted differences were −0.99 percentage points for non-Hispanic Black adults previously treated and, among adults untreated at entry, −2.29 for non-Hispanic Black, +0.42 for Hispanic (not significant), +0.58 for non-Hispanic Asian (not significant), and +3.29 for Other adults. These complete-case estimates are exploratory and based on a selected subset, so primary race and ethnicity conclusions are suggestive.

| Group | N | Unadjusted 5-year remission, % | Adjusted 5-year remission, % (SE) | Adjusted difference vs reference, percentage points (95% CI) |
| --- | --- | --- | --- | --- |
| Previously treated, 1 class | 37,555 | 5.29 | 4.65 (0.12) | Reference |
| Previously treated, 2 classes | 29,609 | 3.56 | 3.62 (0.12) | -1.03 (-1.35 to -0.70) |
| Previously treated, 3 classes | 17,739 | 2.43 | 2.69 (0.14) | -1.95 (-2.32 to -1.58) |
| Previously treated, 4 or more classes | 11,914 | 1.68 | 2.02 (0.17) | -2.63 (-3.04 to -2.22) |
| Previously treated, non-Hispanic White | — | 3.83 | 3.67 (0.08) | Reference |
| Previously treated, non-Hispanic Black | — | 3.60 | 3.61 (0.16) | -0.06 (-0.42 to 0.29) |
| Previously treated, Hispanic | — | 3.08 | 3.52 (0.26) | -0.15 (-0.68 to 0.38) |
| Previously treated, non-Hispanic Asian | — | 3.77 | 3.71 (0.31) | 0.04 (-0.60 to 0.68) |
| Previously treated, Other | — | 4.89 | 4.62 (0.23) | 0.95 (0.48 to 1.42) |
| Untreated at entry, non-Hispanic White | — | 13.78 | 13.75 (0.23) | Reference |
| Untreated at entry, non-Hispanic Black | — | 12.28 | 12.93 (0.43) | -0.83 (-1.77 to 0.12) |
| Untreated at entry, Hispanic | — | 10.27 | 12.10 (0.77) | -1.65 (-3.22 to -0.08) |
| Untreated at entry, non-Hispanic Asian | — | 12.52 | 11.73 (0.77) | -2.02 (-3.60 to -0.44) |
| Untreated at entry, Other | — | 14.49 | 14.59 (0.44) | 0.83 (-0.14 to 1.80) |

5-year estimates are evaluated at 1,890 days (21 x 90-day intervals) and ignore rejected-death censoring. 26.5% of previously treated adults had no class covering the entry date (25,703/96,817).

**Supplementary Table 15.** Medication burden at cohort entry versus immediately before withdrawal among remitters. Entry burden was the number of distinct glucose-lowering classes with a treatment episode recorded at or before entry. Pre-withdrawal burden was the number of distinct classes in the last continuous treatment spell ending before remission. Among all 10,411 remitters, 36.7% changed burden (30.4% increased and 6.3% decreased); median burden rose from 0 to 1 class, mean burden rose from 0.67 to 1.06, and insulin use rose from 13.5% to 25.0%. When insulin was restricted to outpatient sources (outpatient orders and pharmacy dispensed claims, excluding hospital-administered doses), insulin use among all remitters rose from 6.8% at entry to 10.8% immediately before withdrawal (previously treated: 16.8% to 19.9%; untreated at entry: 0.0% to 4.7%), compared with 13.5% to 25.0% using all sources; 56.0% of pre-withdrawal insulin users had only hospital-administered insulin records, so the rise was smaller when hospital doses were excluded. Among 4,188 previously treated remitters, 43.5% changed burden and insulin use rose from 33.5% to 42.4%. Among 6,223 adults untreated at entry, 1,998 later started treatment before remission (1,204 reached 1 class, 492 reached 2, 196 reached 3, and 106 reached 4 or more) and 4,225 remained at 0 classes. Exit-type differences in pre-withdrawal burden are exploratory descriptive findings only.

| **Cohort** | **Entry burden**<br>**(drug classes)** | **0** | **1** | **2** | **3** | **4 or**<br>**more** | **Row N** |
| --- | --- | --- | --- | --- | --- | --- | --- |
| All remitters | 0 | 4225 | 1204 | 492 | 196 | 106 | 6223 |
| All remitters | 1 | 0 | 1579 | 498 | 175 | 91 | 2343 |
| All remitters | 2 | 0 | 355 | 533 | 175 | 117 | 1180 |
| All remitters | 3 | 0 | 109 | 89 | 185 | 90 | 473 |
| All remitters | 4 or more | 0 | 48 | 33 | 19 | 92 | 192 |
| Previously<br>treated at entry | 1 | 0 | 1579 | 498 | 175 | 91 | 2343 |
| Previously<br>treated at entry | 2 | 0 | 355 | 533 | 175 | 117 | 1180 |
| Previously<br>treated at entry | 3 | 0 | 109 | 89 | 185 | 90 | 473 |
| Previously<br>treated at entry | 4 or more | 0 | 48 | 33 | 19 | 92 | 192 |
| Untreated at<br>entry | 0 | 4225 | 1204 | 492 | 196 | 106 | 6223 |

## Supplementary Figures

**Supplementary Figure 1.**
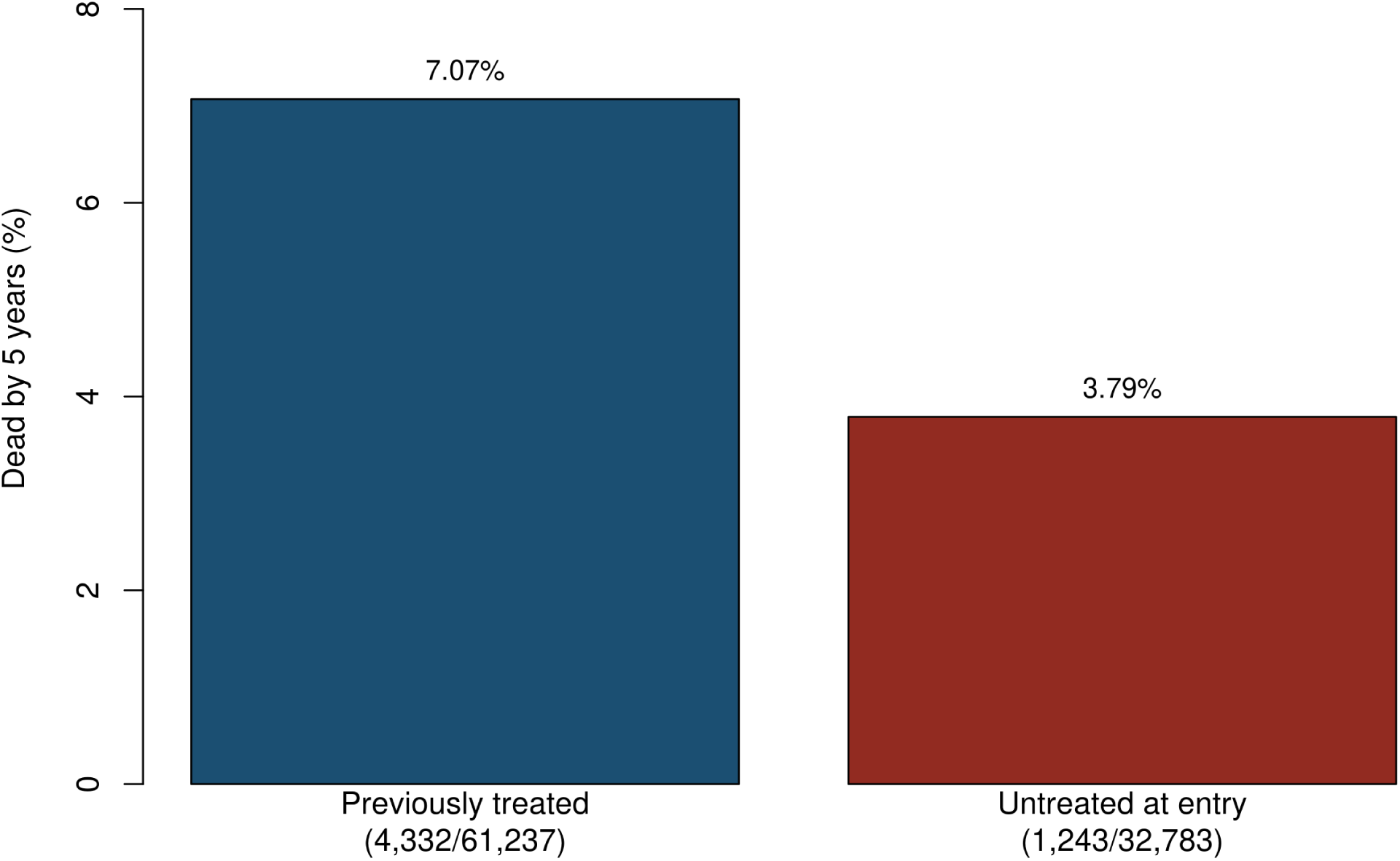
Deaths by 5 years among adults with sufficient follow-up, by treatment history at entry Previously treated: 4,332 deaths among 61,237 adults (7.07%). Untreated at entry: 1,243 deaths among 32,783 adults (3.79%) (Supplementary Table 8). Full glycemic and treatment state occupancy was not estimated in the current analysis.

**Supplementary Figure 2.**
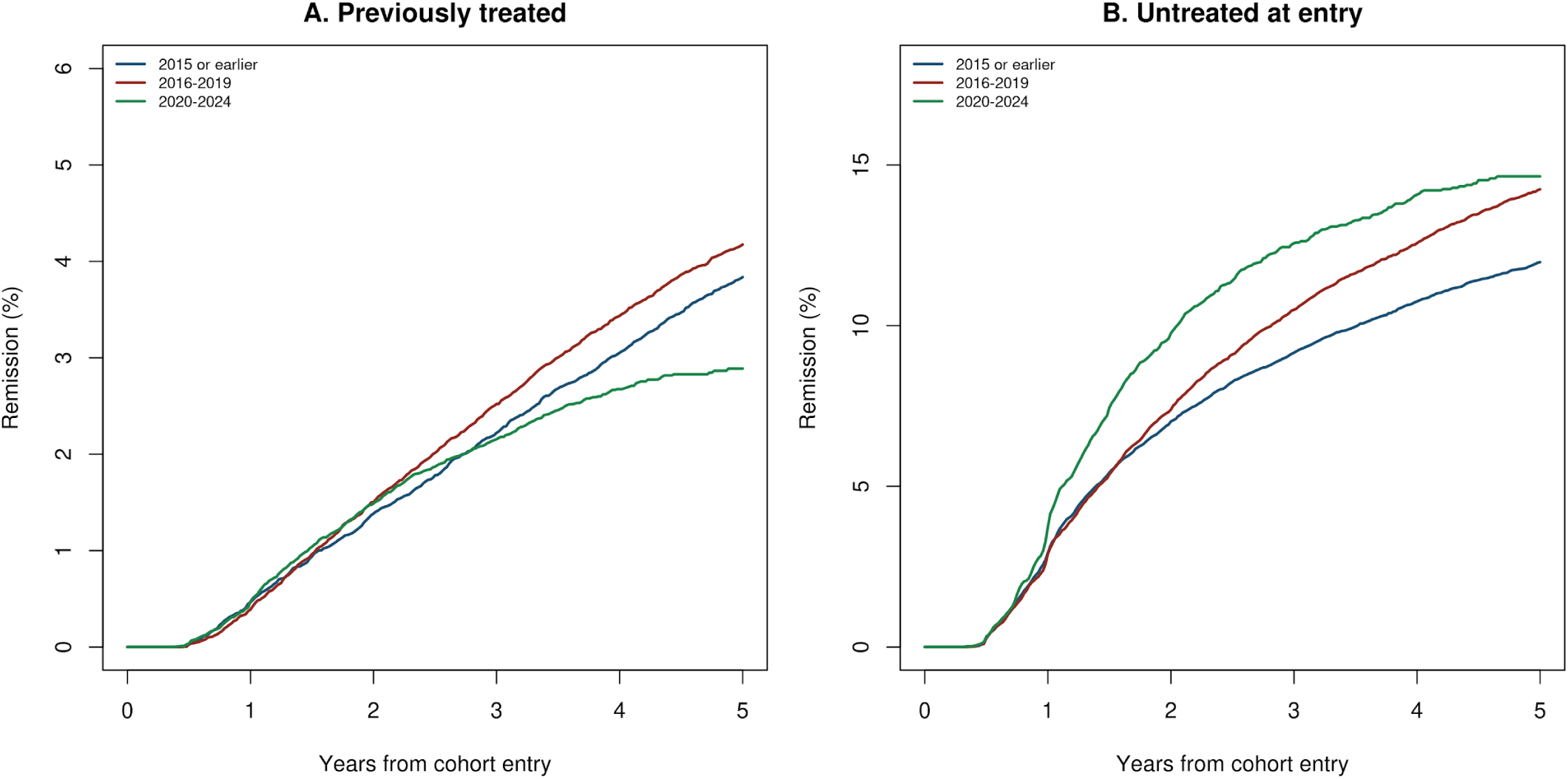
Cumulative incidence of remission by entry period Caution: late entry periods (2020–2024) have short follow-up; 5-year estimates are thin. Five-year estimates are in Supplementary Table 9. Cumulative incidence (Aalen–Johansen, death as a competing event) in previously treated and untreated adults by entry period (2015 or earlier, 2016–2019, 2020–2024). Five-year estimates are in Supplementary Table 9.

## RECORD/STROBE Checklist Form

### Title and abstract

| Item | Recommendation | Location | How addressed |
| --- | --- | --- | --- |
| 1(a) STROBE | Study design in title or abstract | Abstract, Research Design and Methods | Retrospective cohort of adults with laboratory-confirmed type 2 diabetes, followed with competing-risk methods |
| 1(b) STROBE | Informative, balanced summary | Abstract | Structured abstract with objective, design, main estimates with 95% CIs, and conclusions |
| RECORD 1.1 | Type of data; database name | Title; Abstract | Title says “Linked Health Records”; abstract says linked EHR and claims data. Database name (Protégé) not yet in the abstract (gap 1) |
| RECORD 1.2 | Geographic region and timeframe | Abstract | Region: 21 anonymous data-owner values in the delivered data (the vendor attributes 33 contributing systems from a 39-system upstream network). Timeframe not stated because dates are shifted per patient (gap 2) |
| RECORD 1.3 | Linkage between databases stated | Title; Abstract | “Linked” EHR, claims, and death records stated in title and abstract |

### Introduction

| Item | Recommendation | Location | How addressed |
| --- | --- | --- | --- |
| 2 STROBE | Scientific background and rationale | Introduction, paragraphs 1–3 | Consensus definition of remission, trial and real-world estimates, and two gaps: relapse and restart combined in prior studies, and on-treatment control confused with remission |
| 3 STROBE | Objectives and prespecified hypotheses | Introduction, final paragraph | Three objectives: frequency and duration of remission; relapse vs restart as the end of remission; sensitivity of estimates to operational definitions. Descriptive study, no causal hypothesis |

### Methods

| Item | Recommendation | Location | How addressed |
| --- | --- | --- | --- |
| 4 STROBE | Key elements of study design early | Methods, Data Source; Study Population | Retrospective cohort in a linked EHR–claims database; entry at second diagnostic HbA1c |
| 5 STROBE | Setting, locations, dates of recruitment, follow-up, data collection | Methods, Data Source; Supplement S1 | 21 anonymous data-owner values in the delivered data (the vendor attributes 33 contributing systems from a 39-system upstream network); snapshot 4 June 2026; HbA1c follow-up to 30 April 2025; claims 2017–2022; dates shifted per patient |
| 6(a) STROBE | Eligibility, sources and methods of selection, follow-up | Methods, Study Population; Figure 1A | Two HbA1c $\geq 6.5\%$ 90–365 days apart; listed exclusions; previously treated vs untreated at entry; follow-up and censoring rules |
| RECORD 6.1 | Codes and algorithms used to select the population | Methods, Study Population; Supplementary Tables 1 and 1A (code lists) | Laboratory-based case definition (ADA criteria); exclusion and comorbidity code families and HbA1c laboratory codes in the code list |
| RECORD 6.2 | Validation of codes or algorithms | Methods; Limitations | Case definition uses laboratory values per ADA criteria rather than codes alone. No chart validation of the remission or restart phenotype; note-based validation planned as a companion study (gap 3) |
| RECORD 6.3 | Linkage flow diagram with numbers at each stage | Supplement S1 | Vendor token linkage; 4,950,000 patients with EHR data, 4,918,817 with claims and death linkage, 31,183 without. No linkage flow diagram yet (gap 4) |
| 7 STROBE | Define outcomes, exposures, predictors, confounders, effect modifiers | Methods, Remission, Relapse, and Restart; Deaths | Remission, glycemic relapse, treatment restart, end of remission, second and lasting remission, competing death, subgroup variables |
| RECORD 7.1 | Complete list of codes and algorithms | Supplementary Tables 1 and 1A | HbA1c codes and range, nine drug classes, comorbidity ICD-10-CM families, pathway definitions, medication episode algorithm and conventions |
| 8 STROBE | Data sources and measurement for each variable | Methods, Data Source; Medication Records; Supplement S1–S2 | EHR laboratory results, prescriptions, dispensed claims, death file, PCORnet-coded race and ethnicity |
| 9 STROBE | Efforts to address potential sources of bias | Methods, Statistical Analysis; Limitations | Evidence-of-stopping rule with unknown days, competing-risk estimation, death quality checks, and development sample excluded from estimates; testing-intensity weights were not performed in the current analysis |
| 10 STROBE | How study size was arrived at | Methods, Study Population; Figure 1A | All eligible adults in the database; no sample size calculation (descriptive cohort) |
| 11 STROBE | Handling of quantitative variables | Methods; Table 1; Supplementary Table 9 | HbA1c, age, and BMI categorized with stated cut points; confirmation threshold (<6.5%) prespecified; the ≤6.2% confirmation rule was not performed in the current analysis |
| 12(a) STROBE | Statistical methods, including confounding control | Methods, Statistical Analysis | Aalen–Johansen cumulative incidence with death competing; fixed-horizon proportions as a check; descriptive, unadjusted subgroups |
| 12(b) STROBE | Subgroups and interactions | Methods, Statistical Analysis; Table 2 | Treatment history at entry, race and ethnicity, entry HbA1c, age, sex, BMI (descriptive) |
| 12(c) STROBE | Missing data | Methods, Medication Records; Statistical Analysis | Unknown treatment days classified explicitly; BMI analyzed complete-case; zero-unknown-days sensitivity |
| 12(d) STROBE | Loss to follow-up | Methods, Statistical Analysis | Administrative censoring at 30 April 2025; sensitivity censoring at last clinical contact and testing-intensity weights were not performed in the current analysis |
| 12(e) STROBE | Sensitivity analyses | Methods, Statistical Analysis; Supplementary Tables 2 and 4 | Medication rules and population variants; confirmation-rule, testing-weight, censoring, and death-definition sensitivity analyses were not performed in the current analysis |
| RECORD 12.1 | Investigators' access to the database population | Methods, Data Source; Article Information (guarantor) | Authors analyzed the full database population within the governed environment; outputs were aggregate with small cells suppressed |
| RECORD 12.2 | Data cleaning methods | Methods, Medication Records; Supplement S2 | HbA1c range 3.0–20.0% and same-day de-duplication; placeholder dates excluded; dispensed claims only with reversed and rejected claims removed; refinement bridge in Supplementary Table 2 |
| RECORD 12.3 | Linkage level, methods, and quality evaluation | Methods, Data Source; Supplement S1; Deaths | Person-level linkage by the vendor using hashed name, sex, and date-of-birth tokens; linkage quality not independently evaluated except for death records (quality checks, Supplementary Table 3) |

### Results

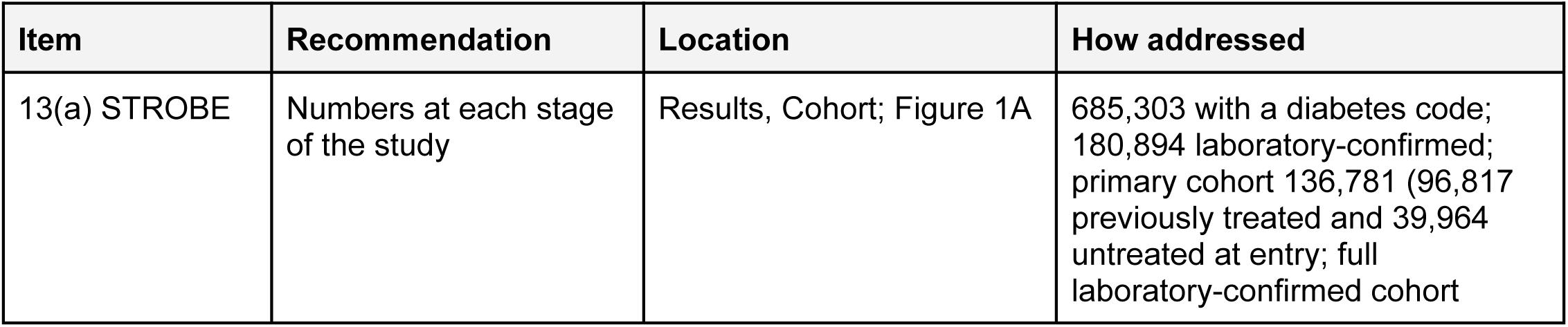
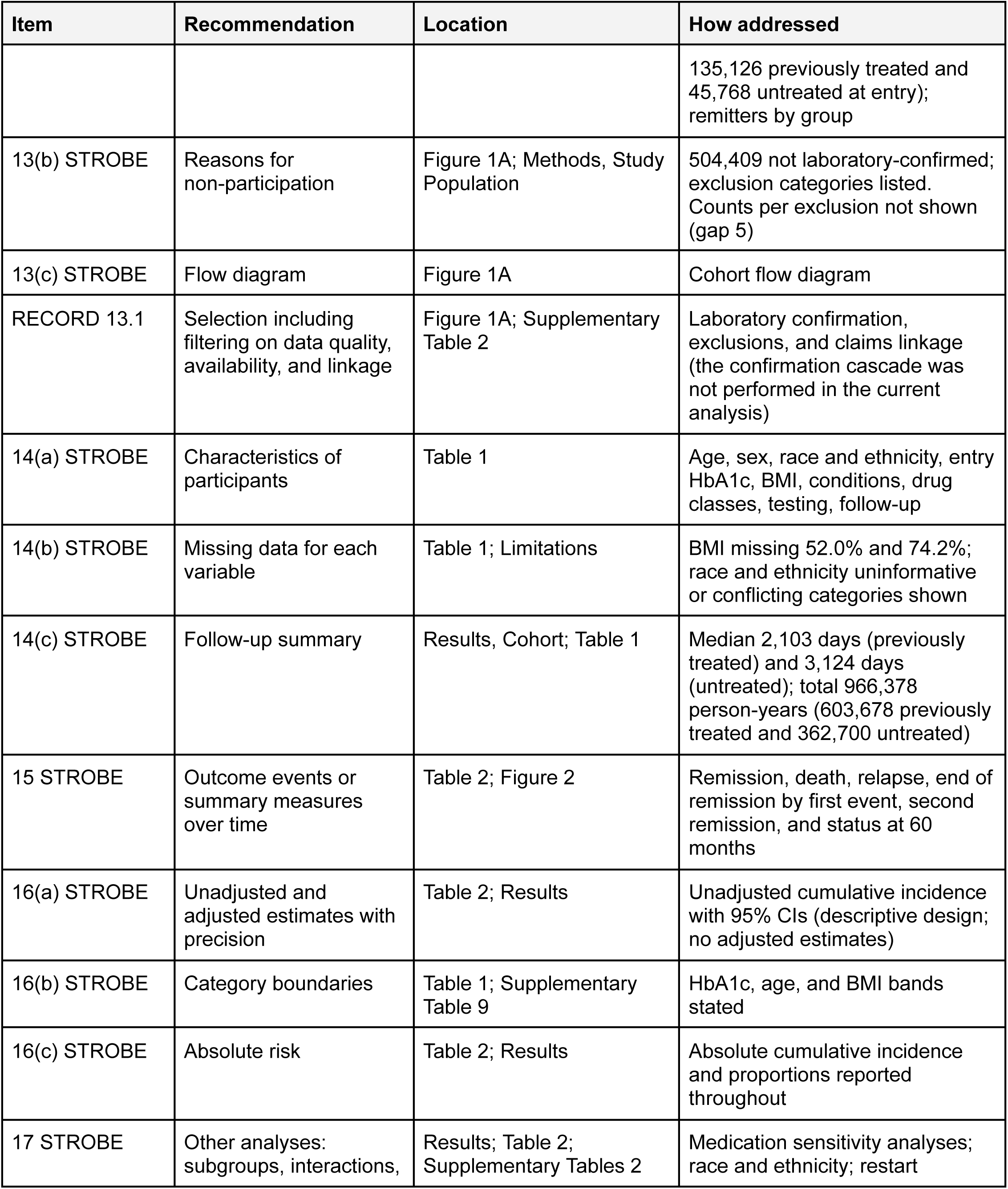
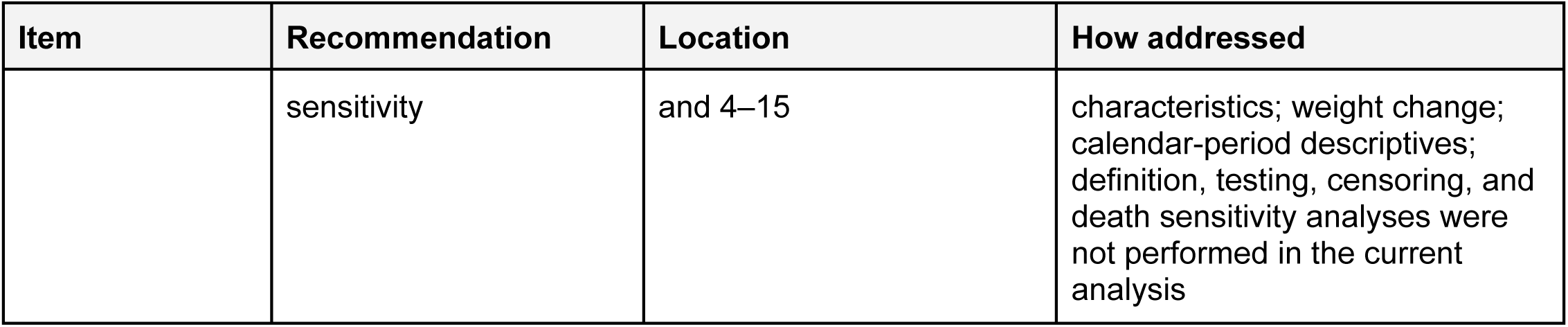

### Discussion

| Item | Recommendation | Location | How addressed |
| --- | --- | --- | --- |
| 18 STROBE | Key results with reference to objectives | Conclusions, paragraph 1 | Frequency, brevity, and how remission ends (restart before relapse) |
| 19 STROBE | Limitations, including bias and imprecision | Conclusions, limitations paragraph | Generalizability, incomplete treatment capture, initial medication definition, death records, date shifting, missing data, unadjusted subgroups, site clustering, direction of bias |
| RECORD 19.1 | Implications of using data not collected for the research question | Conclusions, limitations paragraph; Methods, Development and Confirmation Samples | Misclassification of treatment from open claims and EHR prescriptions; testing-dependent ascertainment; missing BMI; operational rules developed on a separate sample; direction of bias discussed |
| 20 STROBE | Cautious overall interpretation | Conclusions, paragraphs 2–6 | Restart vs relapse interpretation, GLP-1 receptor agonist and weight findings, ascertainment rules, comparison with prior studies, untreated group not a treatment effect |
| 21 STROBE | Generalizability | Conclusions, limitations and trial-comparison paragraphs | Enriched research database, not a population registry; trial results not benchmarks for a prevalent cohort |

### Other information

| Item | Recommendation | Location | How addressed |
| --- | --- | --- | --- |
| 22 STROBE | Funding and role of funders | Article Information, Funding; Duality of Interest | Meta Superintelligence Labs funding; Meta's roles (data license, computing environment, technical support, AI models, draft review without approval rights); C.C.'s employment and authorship; author contracts disclosed |
| RECORD 22.1 | Access to protocol, raw data, or programming code | Article Information, Data Availability | Data under governed access (contact: C.C.); aggregate results, code lists, and analysis code from the corresponding author on request. Protocol availability not yet stated (gap 6) |

## References

1. Riddle MC, Cefalu WT, Evans PH, et al. Consensus report: definition and interpretation of remission in type 2 diabetes. Diabetes Care 2021;44:2438–2444

2. Lean ME, Leslie WS, Barnes AC, et al. Primary care-led weight management for remission of type 2 diabetes (DiRECT): an open-label, cluster-randomised trial. Lancet 2018;391:541–551

3. Lean MEJ, Leslie WS, Barnes AC, et al. Durability of a primary care-led weight-management intervention for remission of type 2 diabetes: 2-year results of the DiRECT open-label, cluster-randomised trial. Lancet Diabetes Endocrinol 2019;7:344–355

4. Lean MEJ, Leslie WS, Barnes AC, et al. 5-year follow-up of the randomised Diabetes Remission Clinical Trial (DiRECT) of continued support for weight loss maintenance in the UK: an extension study. Lancet Diabetes Endocrinol 2024;12:233–246

5. Taheri S, Zaghloul H, Chagoury O, et al. Effect of intensive lifestyle intervention on bodyweight and glycaemia in early type 2 diabetes (DIADEM-I): an open-label, parallel-group, randomised controlled trial. Lancet Diabetes Endocrinol 2020;8:477–489

6. Gregg EW, Chen H, Wagenknecht LE, et al. Association of an intensive lifestyle intervention with remission of type 2 diabetes. JAMA 2012;308:2489–2496

7. Schauer PR, Bhatt DL, Kirwan JP, et al. Bariatric surgery versus intensive medical therapy for diabetes—5-year outcomes. N Engl J Med 2017;376:641–651

8. Mingrone G, Panunzi S, De Gaetano A, et al. Metabolic surgery versus conventional medical therapy in patients with type 2 diabetes: 10-year follow-up of an open-label, single-centre, randomised controlled trial. Lancet 2021;397:293–304

9. Valabhji J, Gorton T, Barron E, et al. Early findings from the NHS Type 2 Diabetes Path to Remission Programme: a prospective evaluation of real-world implementation. Lancet Diabetes Endocrinol 2024;12:653–663

10. Gregg EW, Chen H, Bancks MP, et al. Impact of remission from type 2 diabetes on long-term health outcomes: findings from the Look AHEAD study. Diabetologia 2024;67:459–469

11. Karter AJ, Nundy S, Parker MM, Moffet HH, Huang ES. Incidence of remission in adults with type 2 diabetes: the Diabetes & Aging Study. Diabetes Care 2014;37:3188–3195

12. Sheils NE, Jarvis MS, Bangerter LR, Asch DA, Clark CN. Real-world prevalence of type 2 diabetes remission in a U.S. insured population using a large administrative claims database. Diabetes Spectr 2023;36:211–218

13. Captieux M, Fleetwood K, Kennon B, et al. Epidemiology of type 2 diabetes remission in Scotland in 2019: a cross-sectional population-based study. PLoS Med 2021;18:e1003828

14. Wu H, Yang A, Lau ESH, et al. 1-year weight change after diabetes diagnosis and long-term incidence and sustainability of remission of type 2 diabetes in real-world settings in Hong Kong: an observational cohort study. PLoS Med 2024;21:e1004327

15. Holman N, Wild SH, Khunti K, et al. Incidence and characteristics of remission of type 2 diabetes in England: a cohort study using the National Diabetes Audit. Diabetes Care 2022;45:1151–1161

16. Fadini GP, Giaccari A, Broglio F, et al. Type 2 diabetes remission after initiation of GLP-1 receptor agonists: frequency, characteristics, and outcomes using multiple definitions in an observational study. Lancet Reg Health Eur 2025;59:101499

17. Sherifali DT, Racey ME, Greenway MK, et al. Type 2 diabetes remission: a systematic review and meta-analysis of nonsurgical randomized controlled trials. Diabetes Care 2025;48:2181–2191

18. Khunti K, Papamargaritis D, Aroda VR, et al. Re-evaluating the concept of remission in type 2 diabetes: a call for patient-centric approaches. Lancet Diabetes Endocrinol 2025;13:615–634

19. Thapa B, Schmittdiel JA, Arterburn D, et al. Clinical and demographic characteristics associated with diabetes remission in six integrated health care systems: a retrospective cohort study. Diabetes Care 2025;48:1737–1743

20. Davies MJ, Aroda VR, Collins BS, et al. Management of hyperglycemia in type 2 diabetes, 2022. A consensus report by the American Diabetes Association (ADA) and the European Association for the Study of Diabetes (EASD). Diabetes Care 2022;45:2753–2786

21. Ceriello A, Prattichizzo F, Abdullah ARMS, et al. Causes and consequences of discontinuation of GLP1RAs or tirzepatide. Nat Rev Endocrinol 2026;22:530–542

22. Wilding JPH, Batterham RL, Davies M, et al. Weight regain and cardiometabolic effects after withdrawal of semaglutide: the STEP 1 trial extension. Diabetes Obes Metab 2022;24:1553–1564

23. Goldstein BA, Bhavsar NA, Phelan M, Pencina MJ. Controlling for informed presence bias due to the number of health encounters in an electronic health record. Am J Epidemiol 2016;184:847–855

24. Hripcsak G, Albers DJ. Next-generation phenotyping of electronic health records. J Am Med Inform Assoc 2013;20:117–121

25. Weiskopf NG, Weng C. Methods and dimensions of electronic health record data quality assessment: enabling reuse for clinical research. J Am Med Inform Assoc 2013;20:144–151

26. Benchimol EI, Smeeth L, Guttmann A, et al. The REporting of studies Conducted using Observational Routinely-collected health Data (RECORD) statement. PLoS Med 2015;12:e1001885

27. American Diabetes Association Professional Practice Committee for Diabetes. 2. Diagnosis and classification of diabetes: Standards of Care in Diabetes—2026. Diabetes Care 2026;49(Suppl. 1):S27–S49

28. Pazzagli L, Linder M, Zhang M, et al. Methods for time-varying exposure related problems in pharmacoepidemiology: an overview. Pharmacoepidemiol Drug Saf 2018;27:148–160

29. Aalen OO, Johansen S. An empirical transition matrix for non-homogeneous Markov chains based on censored observations. Scand J Stat 1978;5:141–150

30. Putter H, Fiocco M, Geskus RB. Tutorial in biostatistics: competing risks and multi-state models. Stat Med 2007;26:2389–2430

31. Austin PC, Lee DS, Fine JP. Introduction to the analysis of survival data in the presence of competing risks. Circulation 2016;133:601–609

32. Rohlfing C, Wiedmeyer HM, Little R, et al. Biological variation of glycohemoglobin. Clin Chem 2002;48:1116–1118

33. Sacks DB, Arnold M, Bakris GL, et al. Guidelines and recommendations for laboratory analysis in the diagnosis and management of diabetes mellitus. Diabetes Care 2023;46:e151–e199

34. Korytkowski MT, Muniyappa R, Antinori-Lent K, et al. Management of hyperglycemia in hospitalized adult patients in non-critical care settings: an Endocrine Society clinical practice guideline. J Clin Endocrinol Metab 2022;107:2101–2128

35. Lipska KJ, Ross JS, Miao Y, Shah ND, Lee SJ, Steinman MA. Potential overtreatment of diabetes mellitus in older adults with tight glycemic control. JAMA Intern Med 2015;175:356–362

36. Taylor R, Al-Mrabeh A, Sattar N. Understanding the mechanisms of reversal of type 2 diabetes. Lancet Diabetes Endocrinol 2019;7:726–7

